# Physical interventions to interrupt or reduce the spread of respiratory viruses. Part 1 - Face masks, eye protection and person distancing: systematic review and meta-analysis

**DOI:** 10.1101/2020.03.30.20047217

**Authors:** T Jefferson, MA Jones, L Al-Ansary, GA Bawazeer, EM Beller, J Clark, JM Conly, C Del Mar, E Dooley, E Ferroni, P Glasziou, T Hoffmann, S Thorning, ML van Driel

## Abstract

**OBJECTIVE:** To examine the effectiveness of eye protection, face masks, or person distancing on interrupting or reducing the spread of respiratory viruses.

**DESIGN:** Update of a Cochrane review that included a meta-analysis of observational studies during the SARS outbreak of 2003.

**DATA SOURCES:** Eligible trials from the previous review; search of Cochrane Central Register of Controlled Trials, PubMed, Embase and CINAHL from October 2010 up to 1 April 2020; and forwardand backward citation analysis.

**DATA SELECTION:** Randomised and cluster-randomised trials of people of any age, testing the use ofeye protection, face masks, or person distancing against standard practice, or a similar physical barrier. Outcomes included any acute respiratory illness and its related consequences.

**DATA EXTRACTION AND ANALYSIS:** Six authors independently assessed risk of bias using the Cochrane tool and extracted data. We used a generalised inverse variance method for pooling using a random-effects model and reported results with risk ratios and 95% Confidence Intervals (CI).

**RESULTS:** We included 15 randomised trials investigating the effect of masks (14 trials) in healthcare workers and the general population and of quarantine (1 trial). We found no trials testing eye protection. Compared to no masks there was no reduction of influenza-like illness (ILI) cases (Risk Ratio 0.93, 95%CI 0.83 to 1.05) or influenza (Risk Ratio 0.84, 95%CI 0.61-1.17) for masks in the general population, nor in healthcare workers (Risk Ratio 0.37, 95%CI 0.05 to 2.50). There was no difference between surgical masks and N95 respirators: for ILI (Risk Ratio 0.83, 95%CI 0.63 to 1.08), for influenza (Risk Ratio 1.02, 95%CI 0.73 to 1.43). Harms were poorly reported and limited to discomfort with lower compliance. The only trial testing quarantining workers with household ILI contacts found a reduction in ILI cases, but increased risk of quarantined workers contracting influenza. All trials were conducted during seasonal ILI activity.

**CONCLUSIONS:** Most included trials had poor design, reporting and sparse events. There was insufficient evidence to provide a recommendation on the use of facial barriers without other measures. We found insufficient evidence for a difference between surgical masks and N95 respirators and limited evidence to support effectiveness of quarantine. Based on observational evidence from the previous SARS epidemic included in the previous version of our Cochrane review we recommend the use of masks combined with other measures.

## Introduction

Epidemic and pandemic respiratory infections pose a serious threat to people worldwide. Recent pandemics were the H1N1 influenza caused by the H1N1pdm09 virus in 2009 and the current Coronavirus Disease-2019 - COVID-19 - caused by SARS-CoV-2; recent epidemics of note were the Severe Acute Respiratory Syndrome (SARS) in 2003 and the Middle East Respiratory Syndrome (MERS), which began in 2012. Even non-epidemic acute respiratory infections (ARIs) place a huge burden on the healthcare systems of countries and are a prominent cause of morbidity. ^1^In addition, ARIs are often pre-cursors to lower respiratory tract infections (e.g. pneumonia) caused by bacterial pathogens which cause millions of deaths worldwide.

Epidemics and pandemics are more likely following antigenic change in the virus or transmission from animals (domestic or wild) when there is no natural human immunity.^2^ High viral load, high levels of transmissibility, susceptible populations and symptomatic patients are considered to be the drivers of such epidemics and pandemics. ^3^ Most single intervention measures (such as the use of vaccines or antivirals) will be insufficient to contain the spread of influenza^4 5^; but combinations of measures may reduce the reproduction number below 1. For some infectious agents, there are no licensed interventions. Stopping the spread of the virus from person to person via a combination of social and physical interventions may be the only option to reduce the spread of outbreaks.

Physical interventions, such as the use of masks and person distancing measures, might prevent the spread of virus transmitted by aerosols or large droplets from infected to susceptible people. Use of hand hygiene, gloves, and protective gowns can also prevent the spread by limiting the transfer of viral particles onto and from surfaces. Such interventions were emphasized in WHO’s latest Global Influenza Strategy 2019 – 2030^6^ and can have several possible advantages over other methods of suppressing ARI outbreaks: they can be instituted rapidly and may be independent of any specific type of infective agent including novel viruses.

The benefits of physical interventions are self-evident and have been confirmed by evidence included in three previous reviews. ^7 8^ Given the global importance of interrupting viral transmission in the current COVID-19 pandemic, up-to-date estimates of their effectiveness are necessary to inform planning, decision-making, and policy. In this review we concentrate on the evidence for use of eye protection or masks and the effects of person distancing. The next part of this review will include evidence for all other physical interventions.

## Methods

### Inclusion criteria

We included randomised controlled trials (RCT) and cluster-randomised controlled trials (C-RCT) including people of any age that tested the use of face masks (i.e. surgical or medical masks and N95 respirators), eye protection, or person distancing against standard practice, or a similar physical barrier, or compared any of these interventions. We only included studies that reported a measure of acute respiratory illness – such as influenza-like illness, influenza, or respiratory infections – and/or its consequences (e.g. days off work, complications, hospitalisation and death, if clearly reported as consequences of the respiratory illness). We also included relevant studies from the previous versions of this review.^7-9^

### Search strategy

We identified RCTs and C-RCTs studying effectiveness of eye protection (any purposed device excluding simple eyeglasses), masks (defined as any type of facial mask), and person distancing from our 2011 review. ^8^ These earlier studies were analysed using word frequency to create a new search string that was run in PubMed. ^10^ This search string was converted using the Polyglot Search Translator^11^ and run in the following additional databases; the Cochrane Central Register of Controlled Trials, Embase and CINAHL. The search covered the dates October 2010 to 9 March 2020. Search strings for all databases are available in the appendices (Appendix 1). A backwards and forward citation analysis, using Scopus, was conducted on all new studies retrieved. Search and citation analysis results were screened using the RobotSearch tool to remove all obvious non-RCTs. ^12^ Three authors (JC, MJ and ST) independently reviewed the titles and abstracts of the identified studies to assess eligibility for inclusion. Discrepancies were resolved by consensus.

### Risk of bias assessment

Risk of bias was assessed by three author pairs independently (TJ, EB, LA, GB, MJ, EF) for the method of random sequence generation and allocation concealment (selection bias), blinding of participants and personnel (performance bias), blinding of outcome assessment (detection bias), outcome reporting (attrition bias), and selective reporting (reporting bias). We used the Cochrane risk of bias tool. ^13^ For each item risk was either ‘high’, ‘low’ or ‘unclear’. Low risk of bias for the method of random sequence generation indicates that the method was well-described and is likely to produce balanced and truly random groups; for allocation concealment that the next treatment allocation was not known to participant/cluster or treating staff until after consent to join the study; for blinding of participants and personnel that the method is likely to maintain blinding throughout the study; for blinding of outcome assessors that all assessing outcomes were unaware of treatment allocation; for outcome reporting that participant attrition through the study is reported and reasons for loss are appropriately described; and for selective reporting that all likely planned and collected outcomes have been reported.

### Data extraction and analysis

Six authors (TJ, EB, LA, GB, MJ, EF) independently extracted data in pairs. Discrepancies were resolved by consensus. Descriptions of the interventions were extracted using the Template for Intervention Description and Replication (TIDieR) template. ^14^ We entered outcome data in RevMan software and used a generalised inverse variance random effects method for pooling. The effect estimate was expressed as a risk ratio with 95% confidence interval. We calculated the I^2^ statistic for each pooled estimate to assess statistical heterogeneity. ^15^ For studies that could not be pooled we report the effect estimates as reported by the study authors. We conducted a subgroup analysis for interventions aimed at protecting health care workers.

### Differences between 2009 review and current review

The 2009 review included both randomised trials, cluster randomised trials, and observational studies. This update excluded the latter. This update was also split into two parts, according to intervention categories. This Part 1 is focussed on face masks, eye protection, and person distancing; Part 2 will address all other categories of physical interventions.

## Results

### Results of the search

The updated search yielded 2468 references after removal of duplicates of which 2345 were excluded. A further 42 were excluded after review of the full text paper. Backwards and forward citation analysis identified a further 20 studies resulting in a total of 101 papers testing a range of barrier interventions aimed at interrupting the spread of respiratory viruses. For this part 1 of our review we included 15 RCT/c-RCTs, including 5 trials from the 2011 review^16-20^ (see Figure 1).

**Figure 1:**
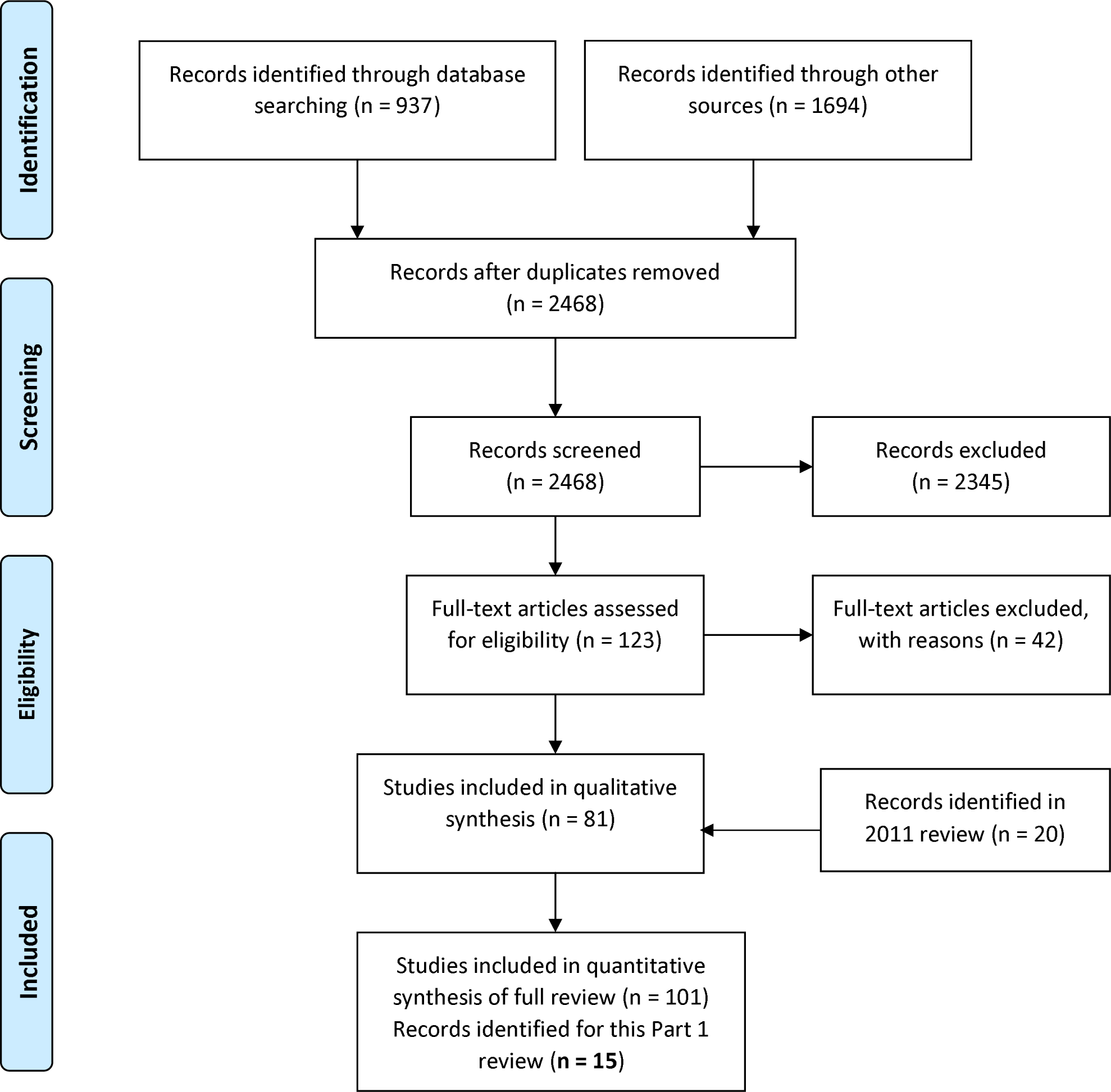
PRISMA Flow Diagram.

### Risk of bias

Reporting of sequence generation, allocation concealment (particularly in cluster-randomised trials) and blinding of outcome assessment was poor, leading to an unclear risk of bias judgement for 30% of studies (see Figure 2). The majority of studies were unblinded due to the nature of the masks, or insufficient information (70% of studies with high or unclear risk of bias). Only one study was blinded to staff. ^21^ For the remainder of the unblinded studies at low risk of bias, this was due to them having objective outcomes that were unlikely to be affected by unblinding. More than 80% of studies had no evidence of serious attrition and described reasons for losses to follow-up well. 70% of studies had no evidence of selective outcome reporting. One study had what appeared to be selective testing or reporting of viral tests, another had selective reporting of non-viral isolates and changes during the study that made planned outcomes unclear. The remainder of the studies had unclear risk of bias for this domain due to insufficient information reported.

**Figure 2.**
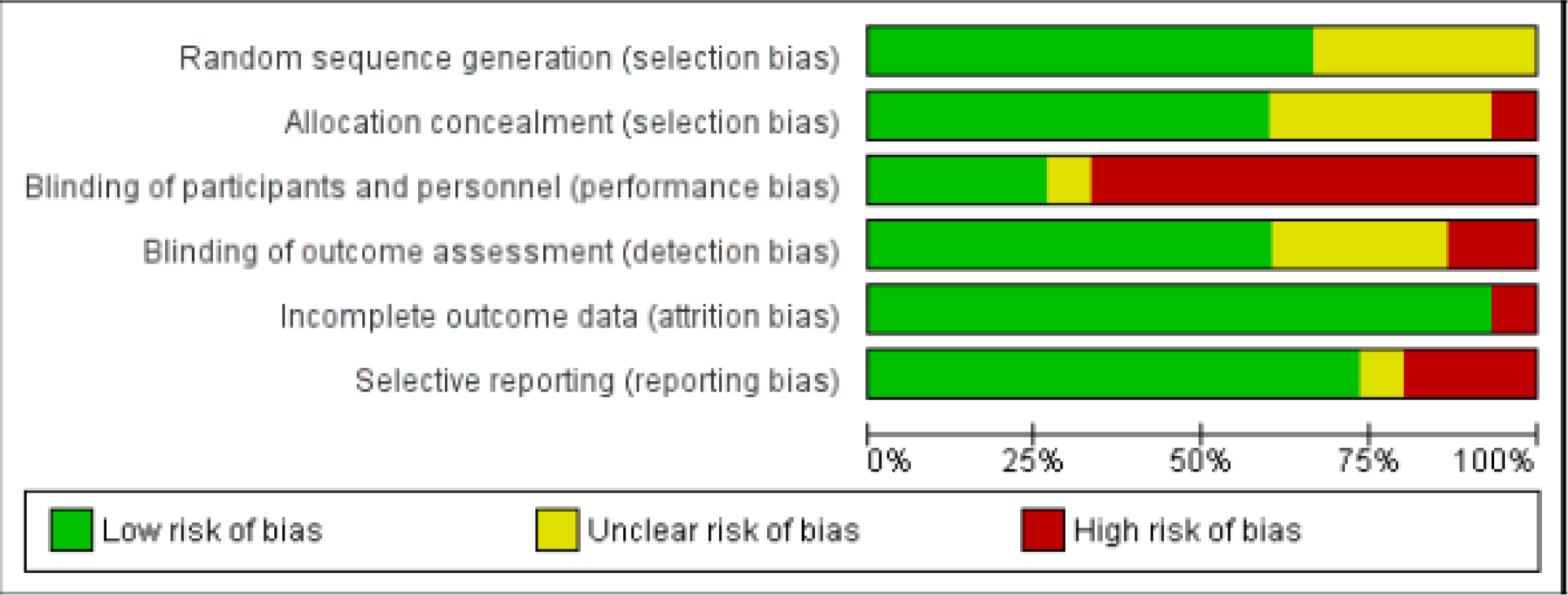
Risk of Bias in included studies - summary bar chart

### Eye protection

We found no trials on the use of eye protection as a single intervention.

### Masks

Nine trials compared masks with no masks. ^16-18 21-26^ Two of these studies included health care workers^18 25^ and 7 others included people living in the community. All trials were conducted in non-pandemic settings. A description of the interventions is presented in Table 1. Included trials are described in Table 2. Pooling of all nine trials did not show a statistically significant reduction of ILI cases (Risk Ratio 0.93, 95%CI 0.83 to 1.05) or laboratory-confirmed influenza cases (Risk Ratio 0.84, 95%CI 0.61-1.17) in the group wearing a mask compared to those not wearing a mask (see Figure 3a). Eight-seven percent of the weight of this analysis is carried by two studies from the same first author. ^16 22^ A separate analysis of the two trials in healthcare workers also failed to show a statistically significant difference between the mask and no mask groups (Risk Ratio 0.37, 95%CI 0.05 to 2.50).

**Table 1.**
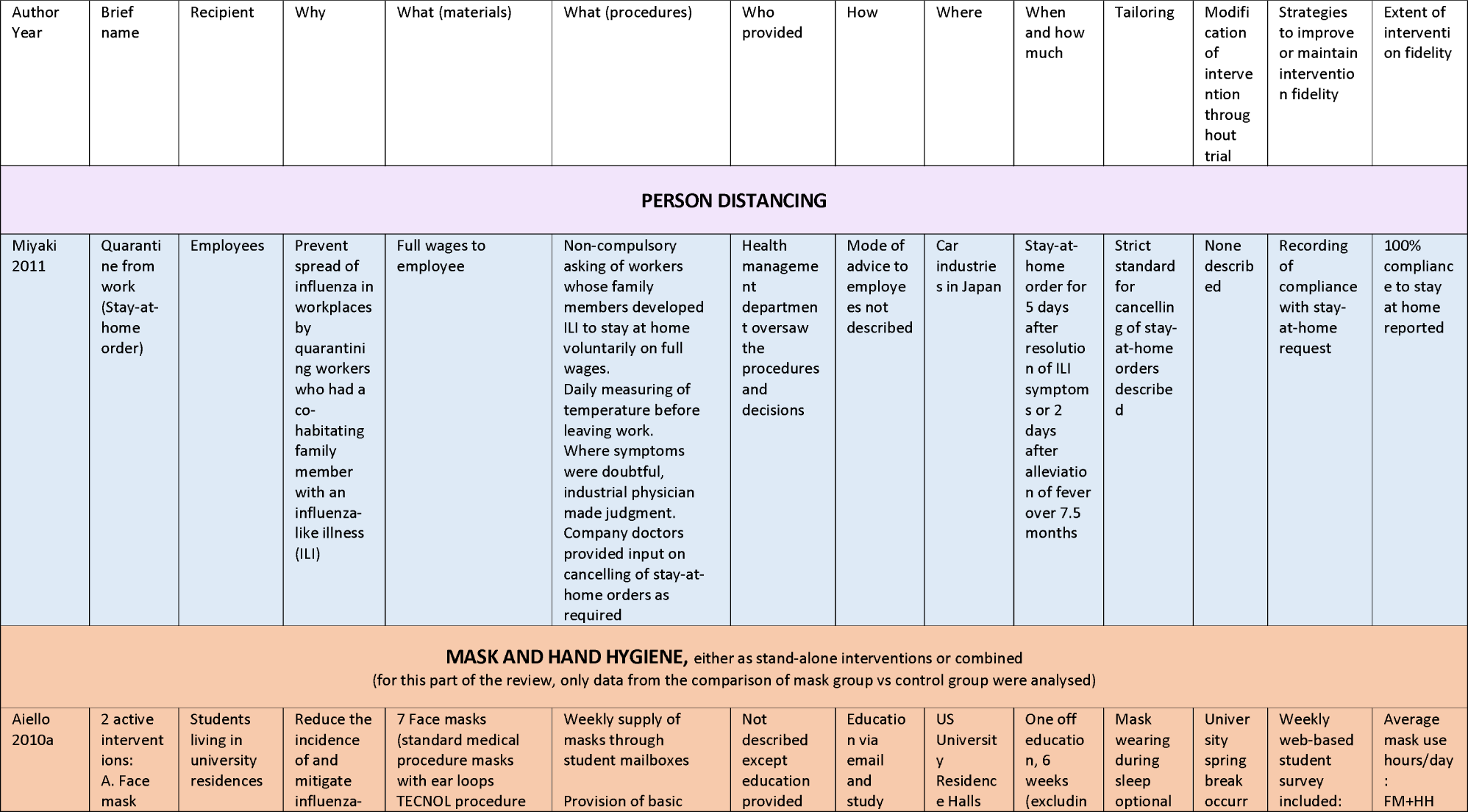
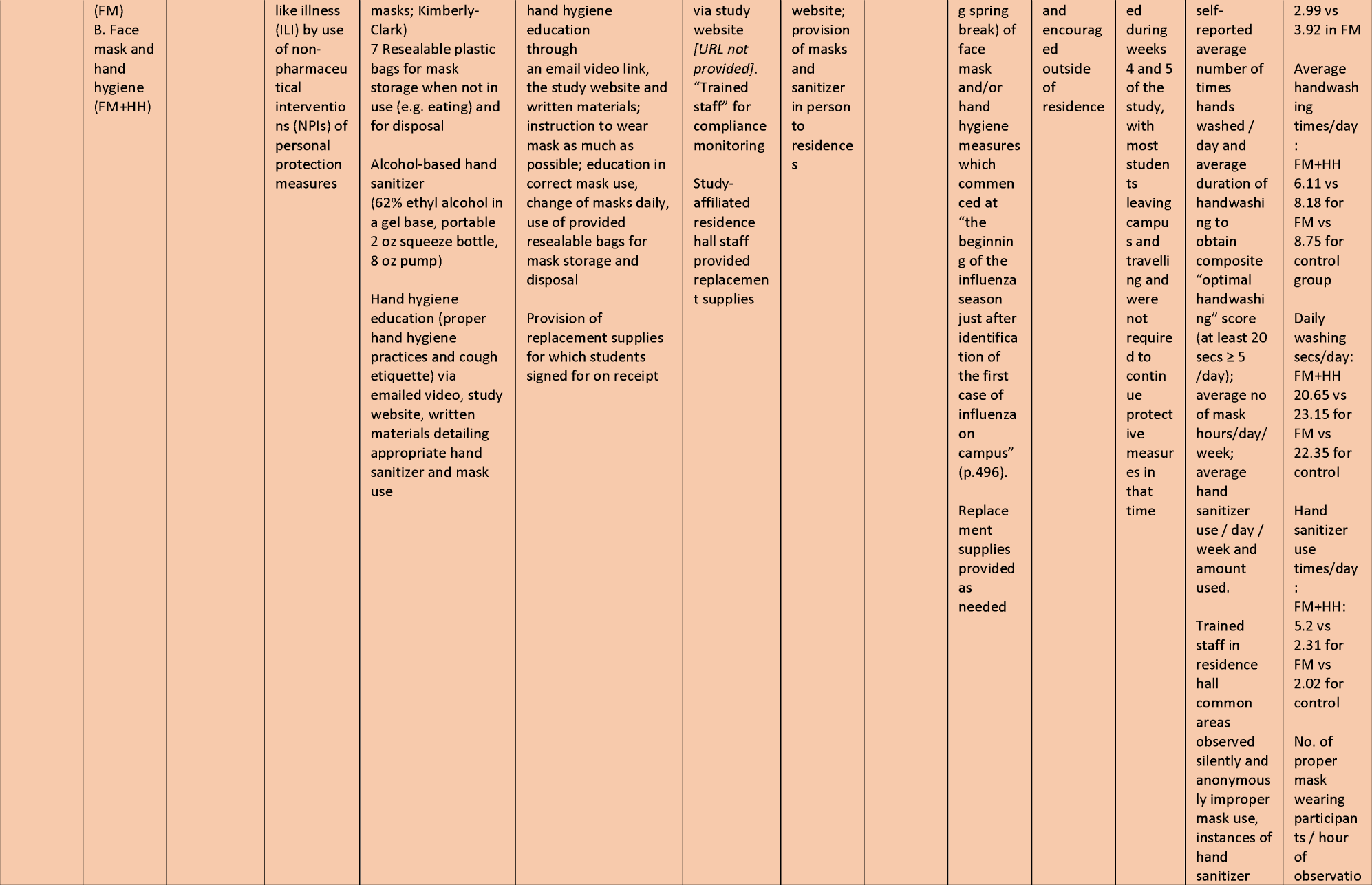
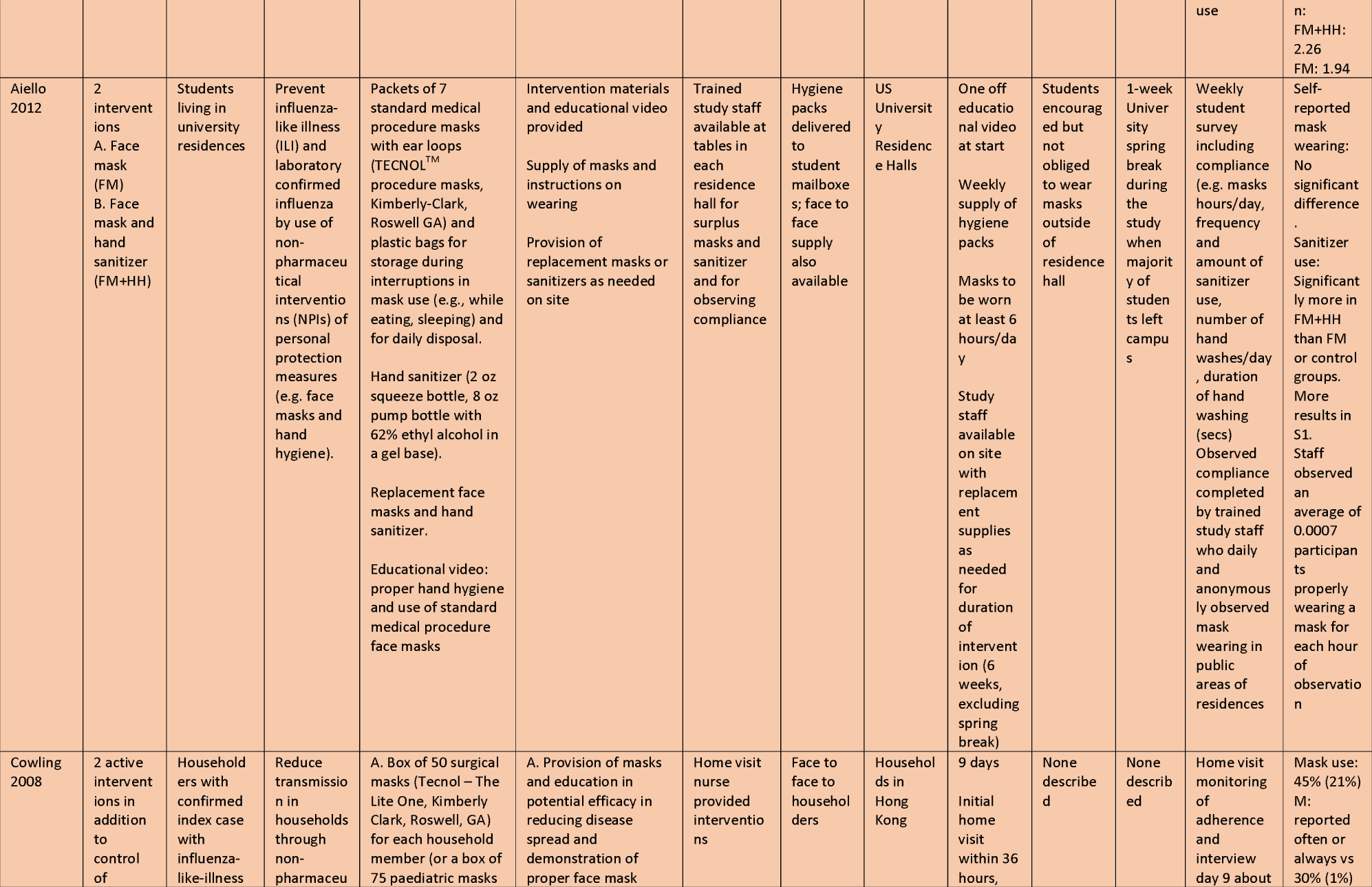
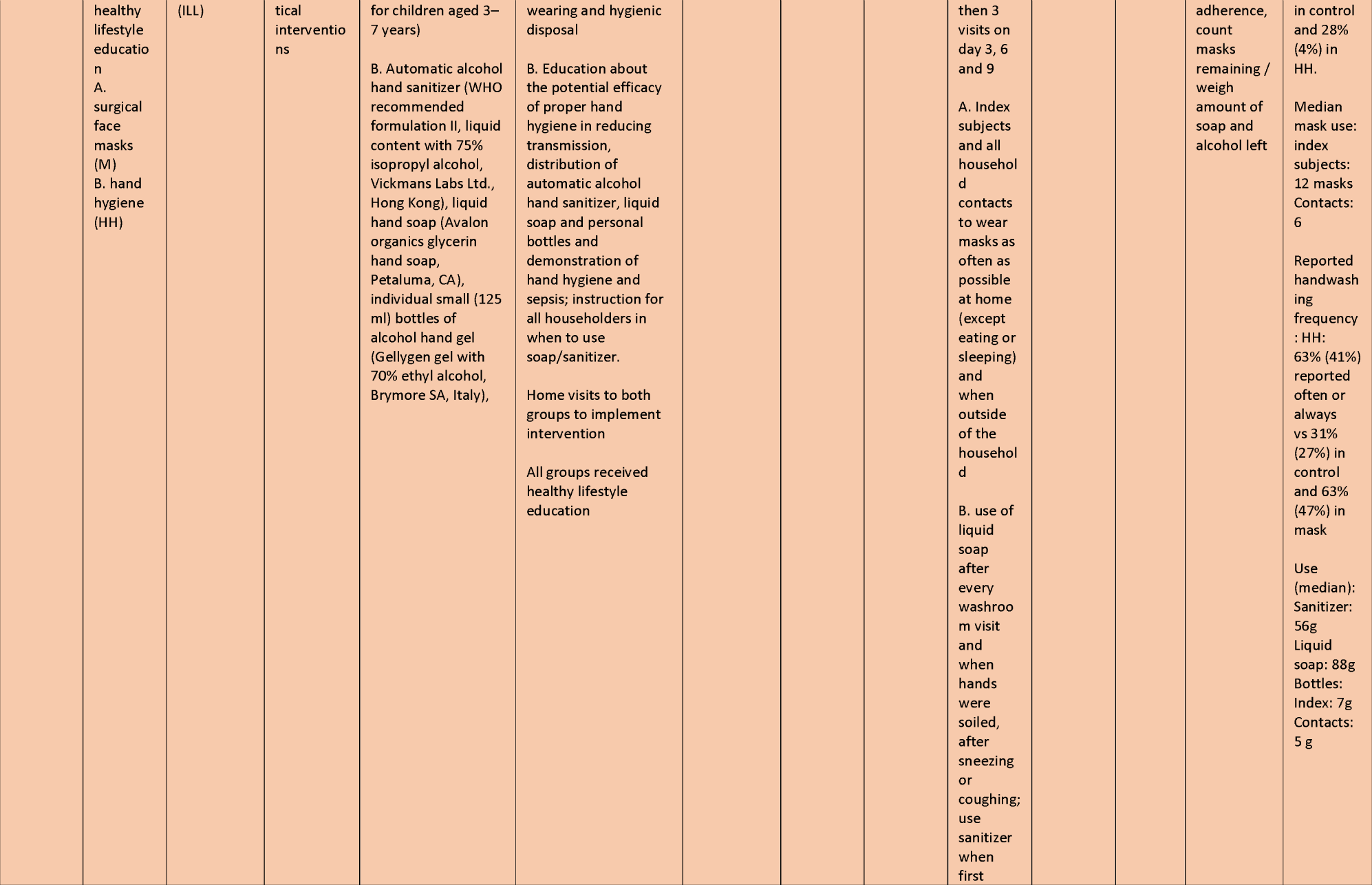
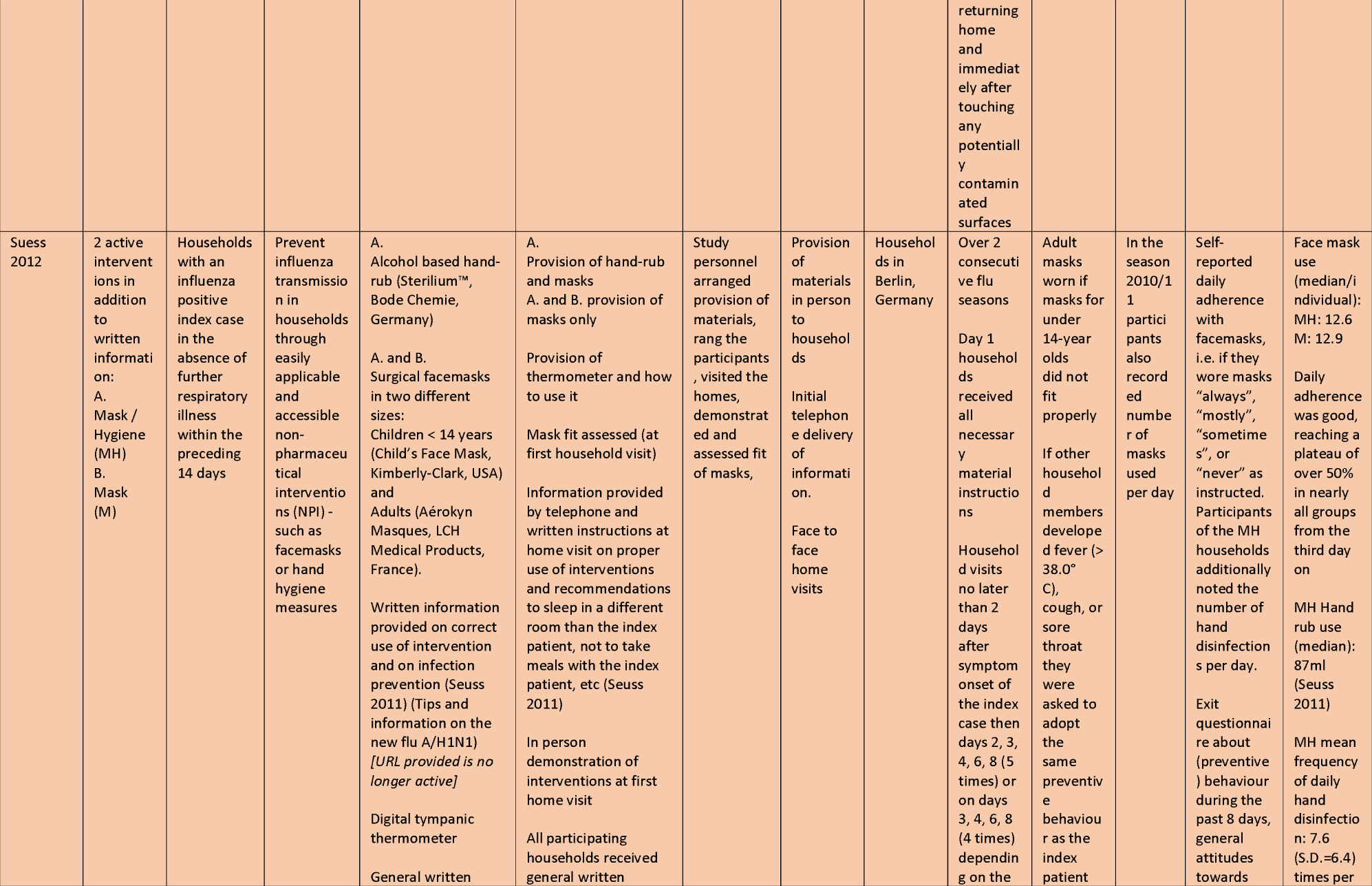
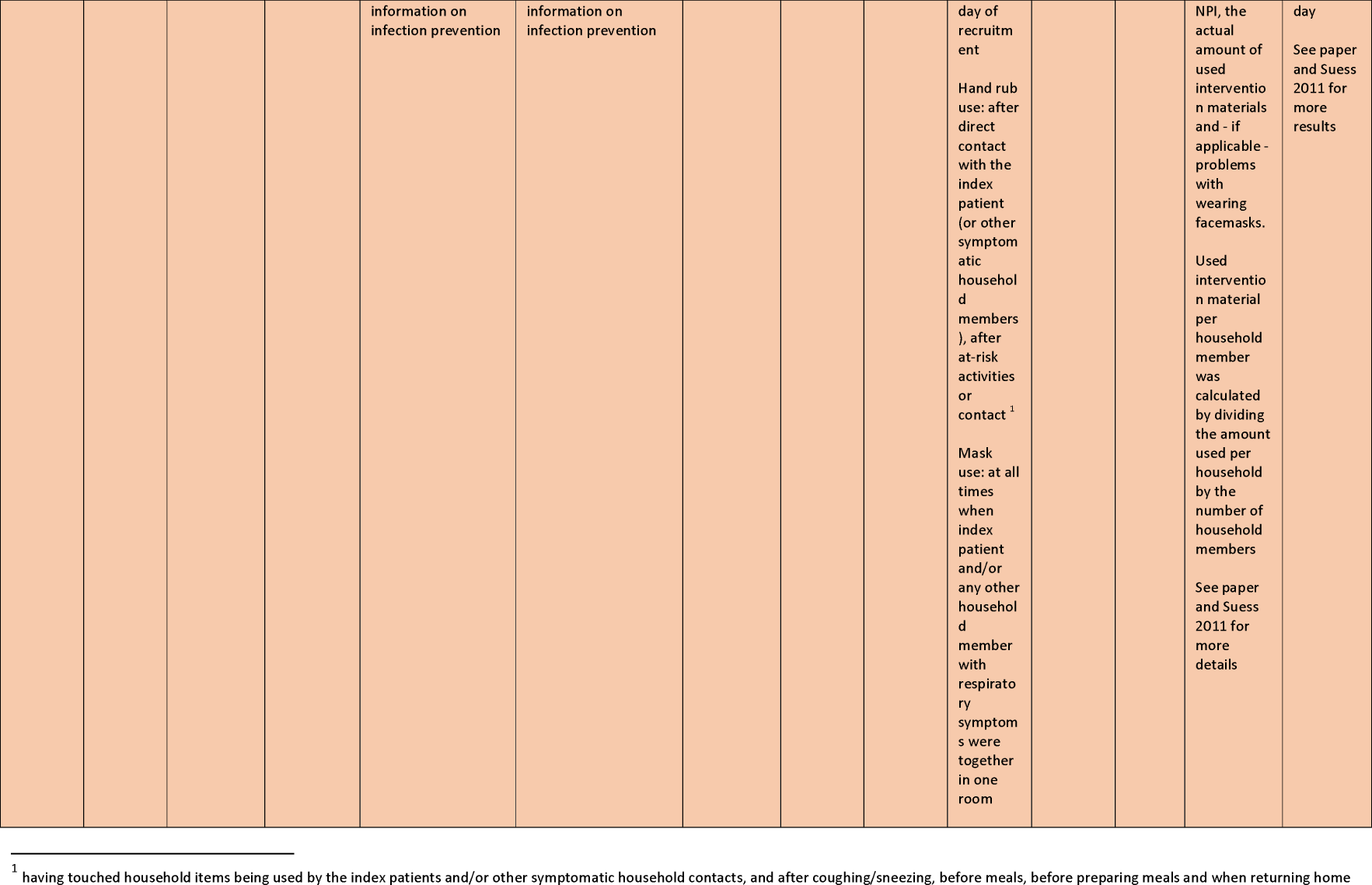
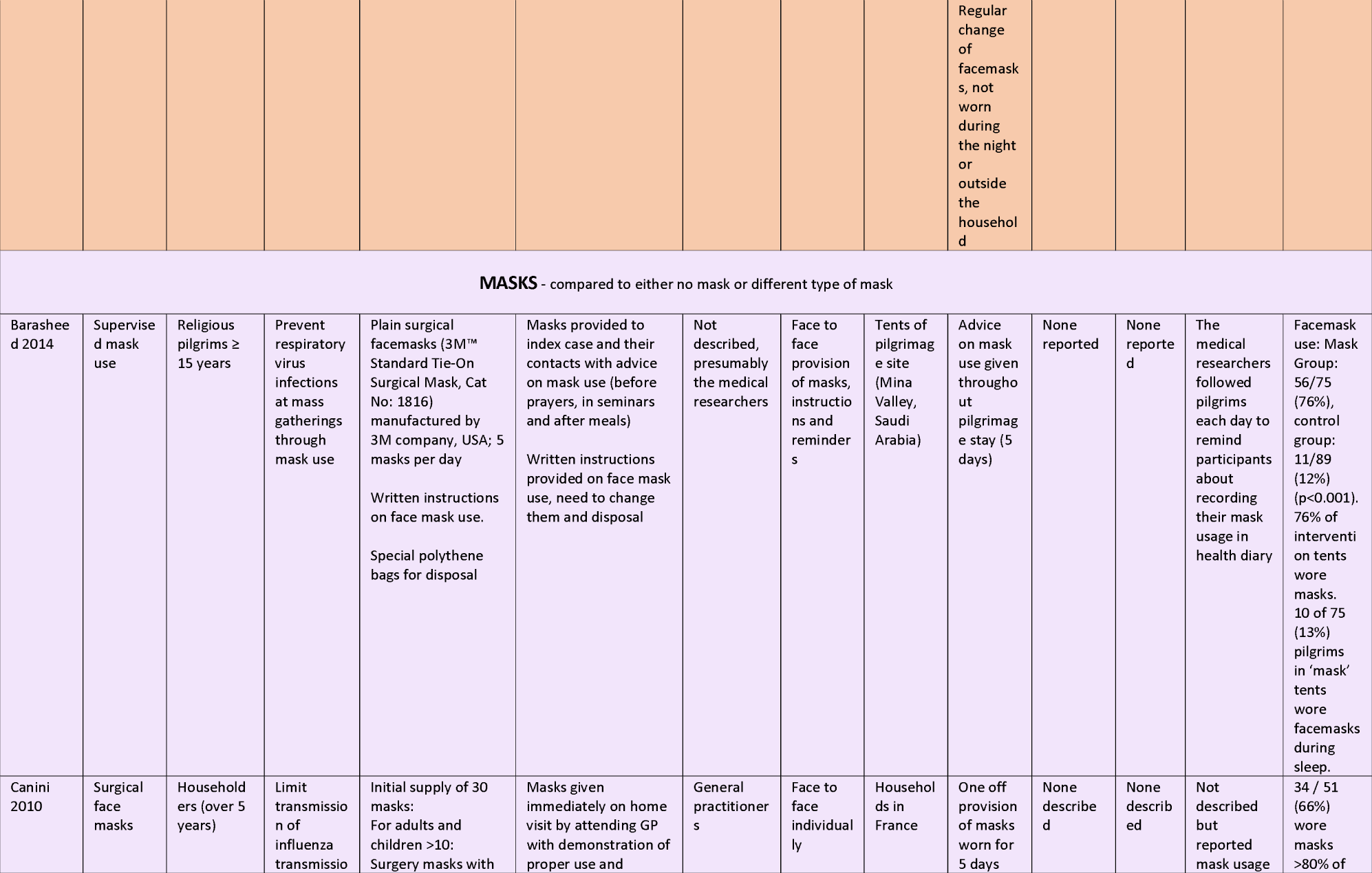
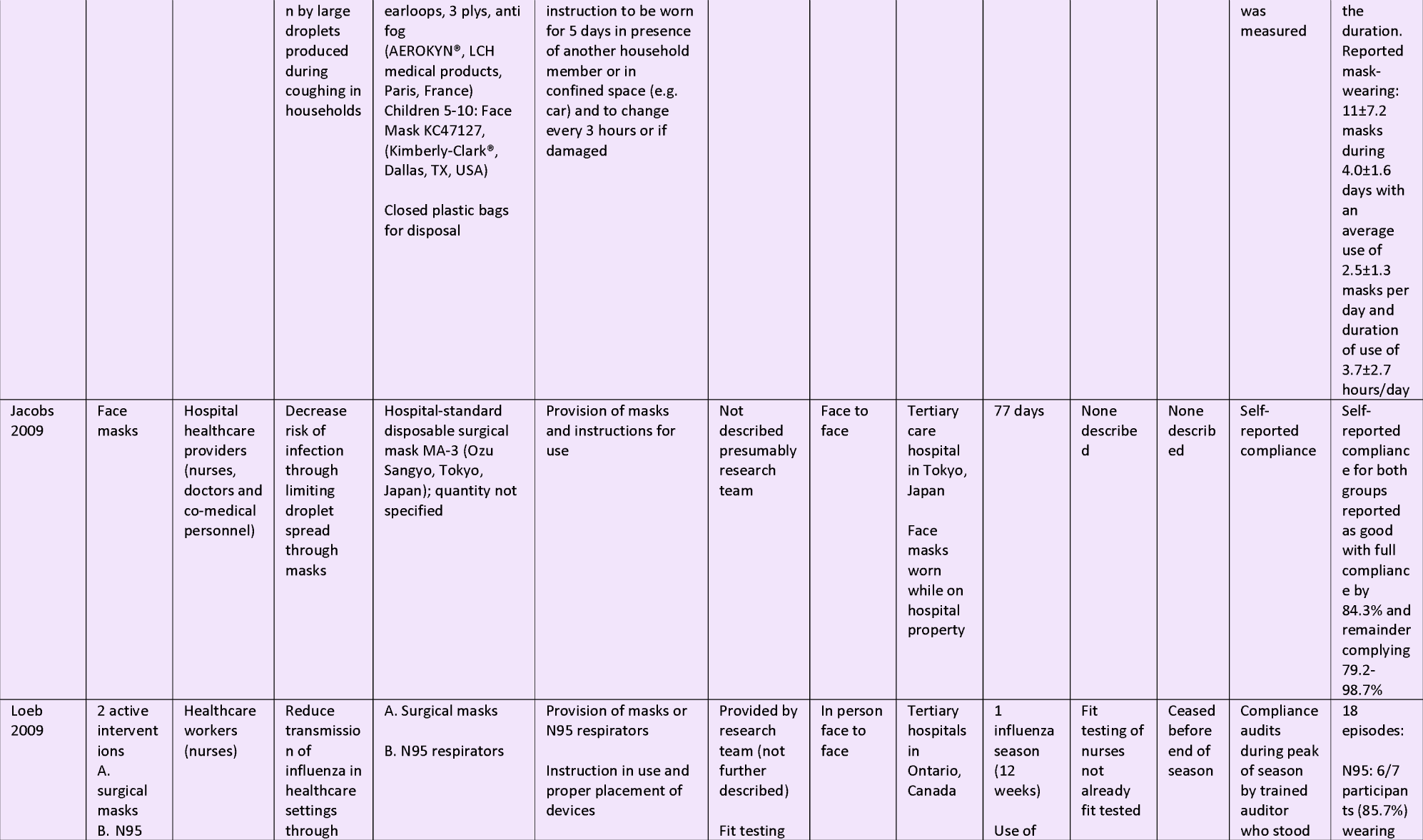
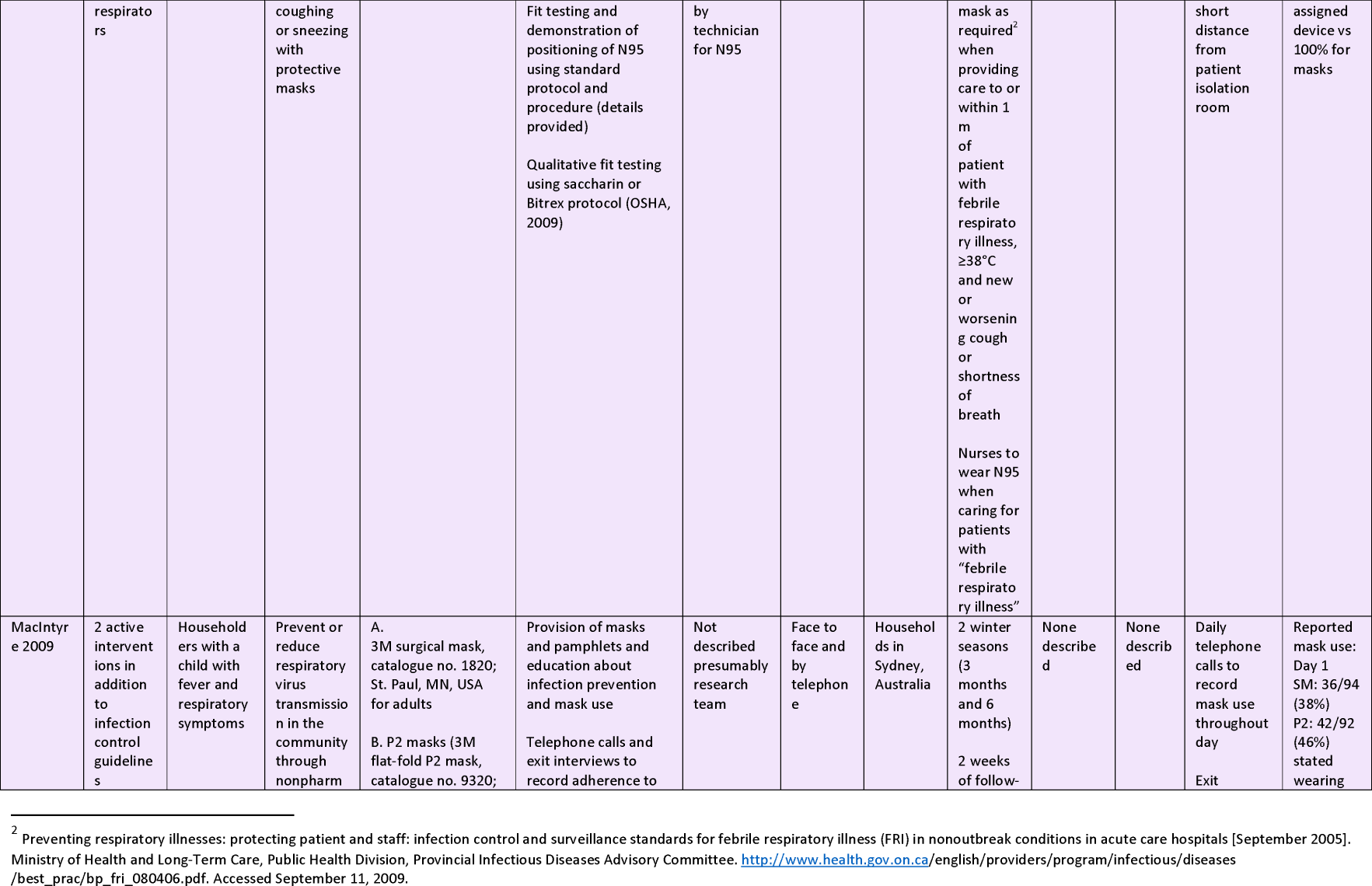
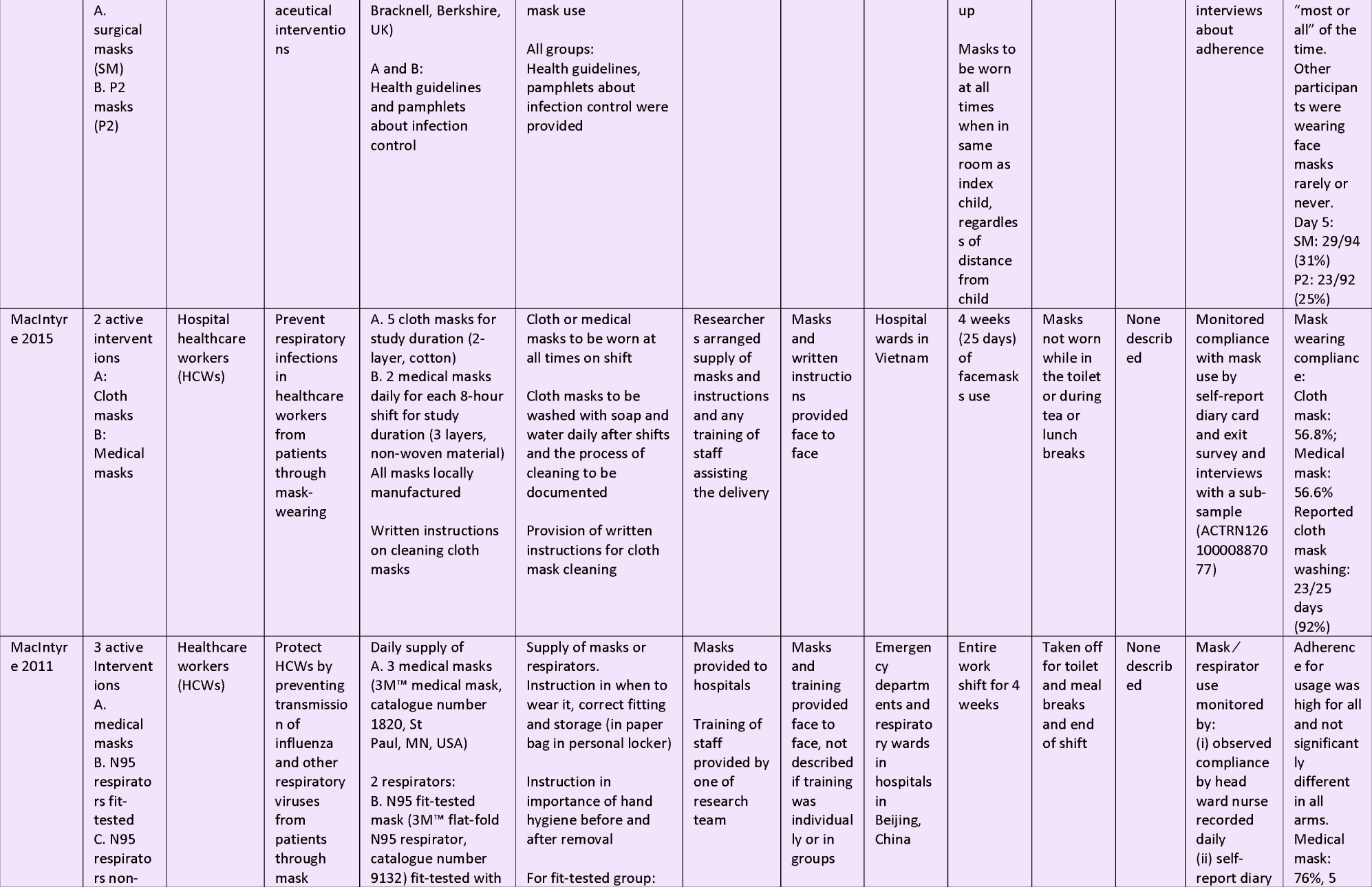
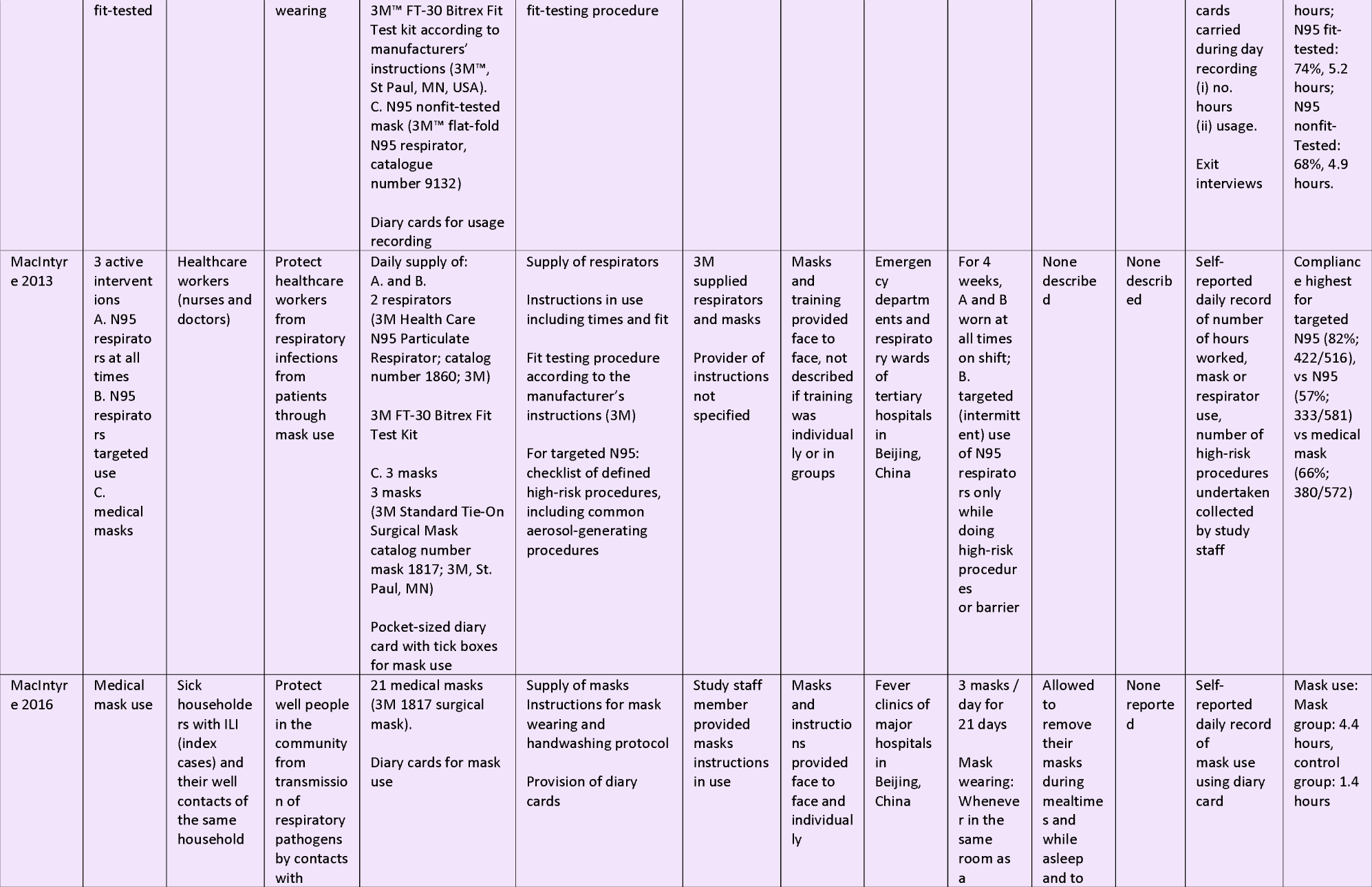
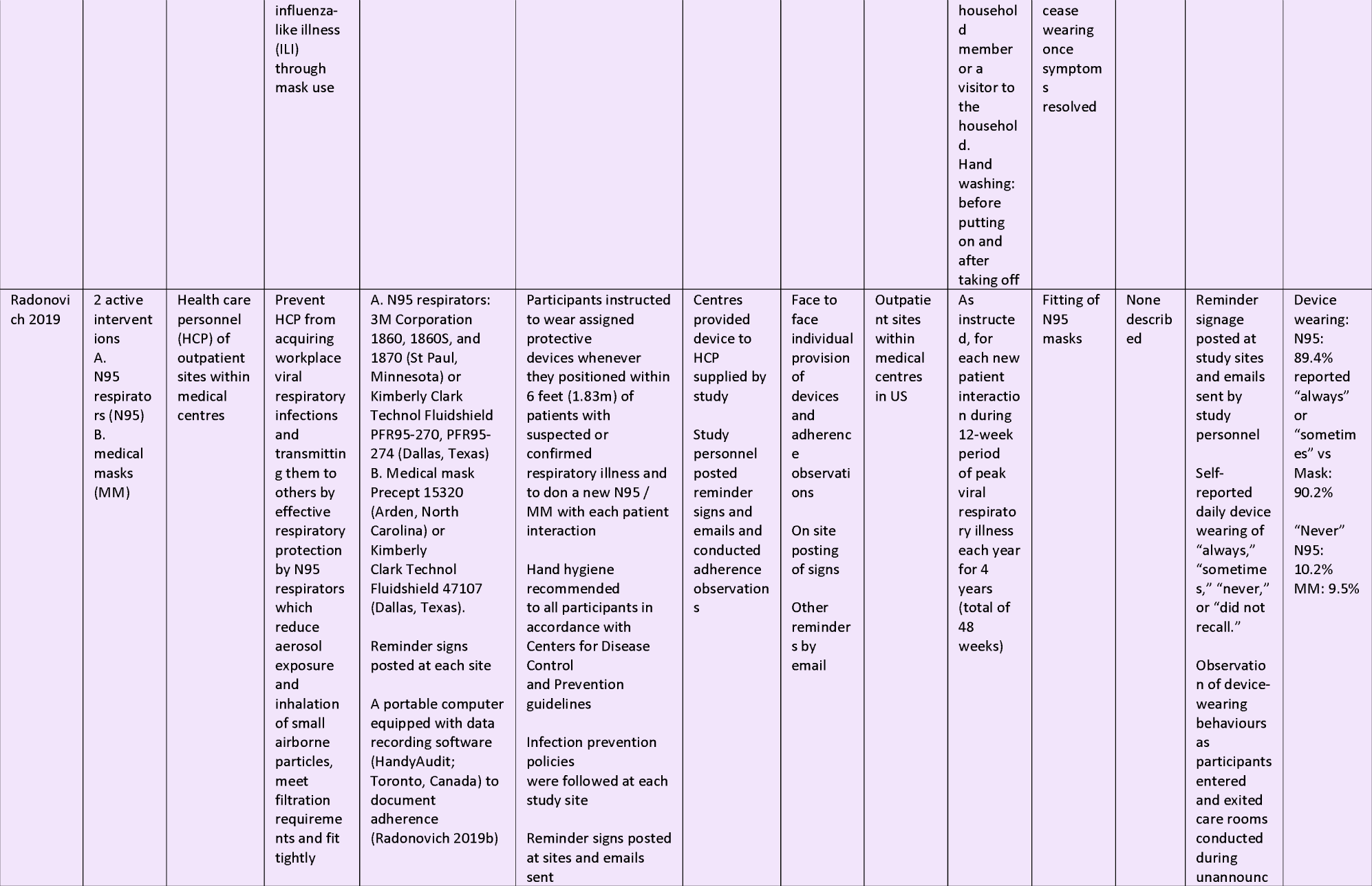
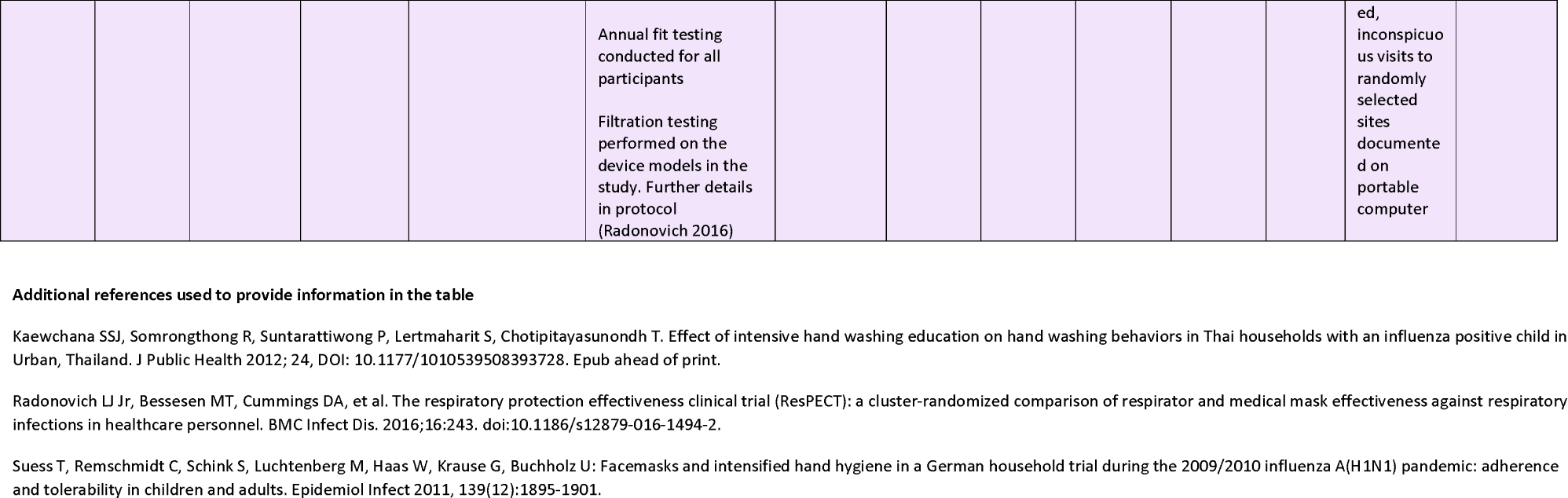
Description of interventions in included studies, using the items from the Template for Intervention Description and Replication (TIDieR) checklist

**Table 2:**
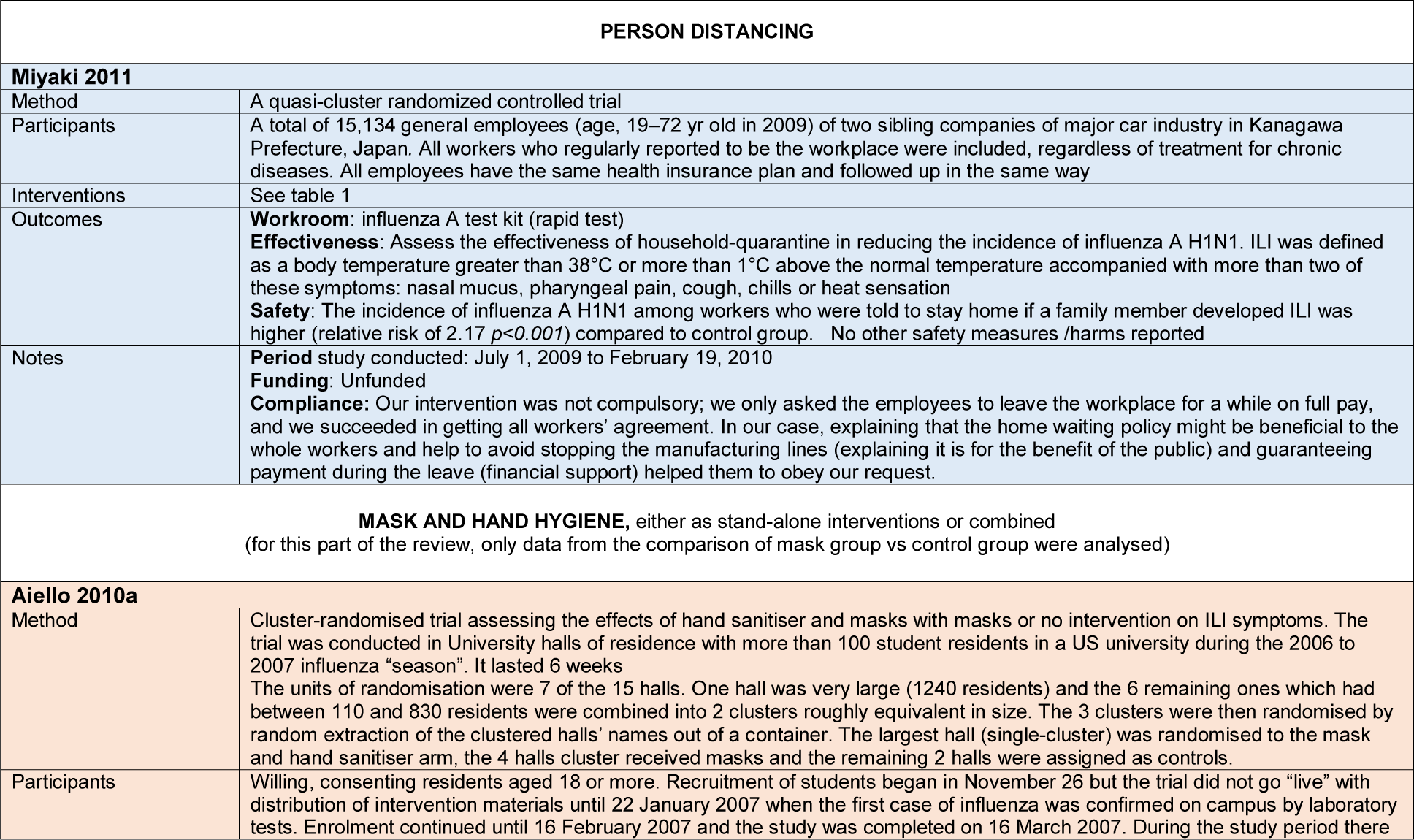
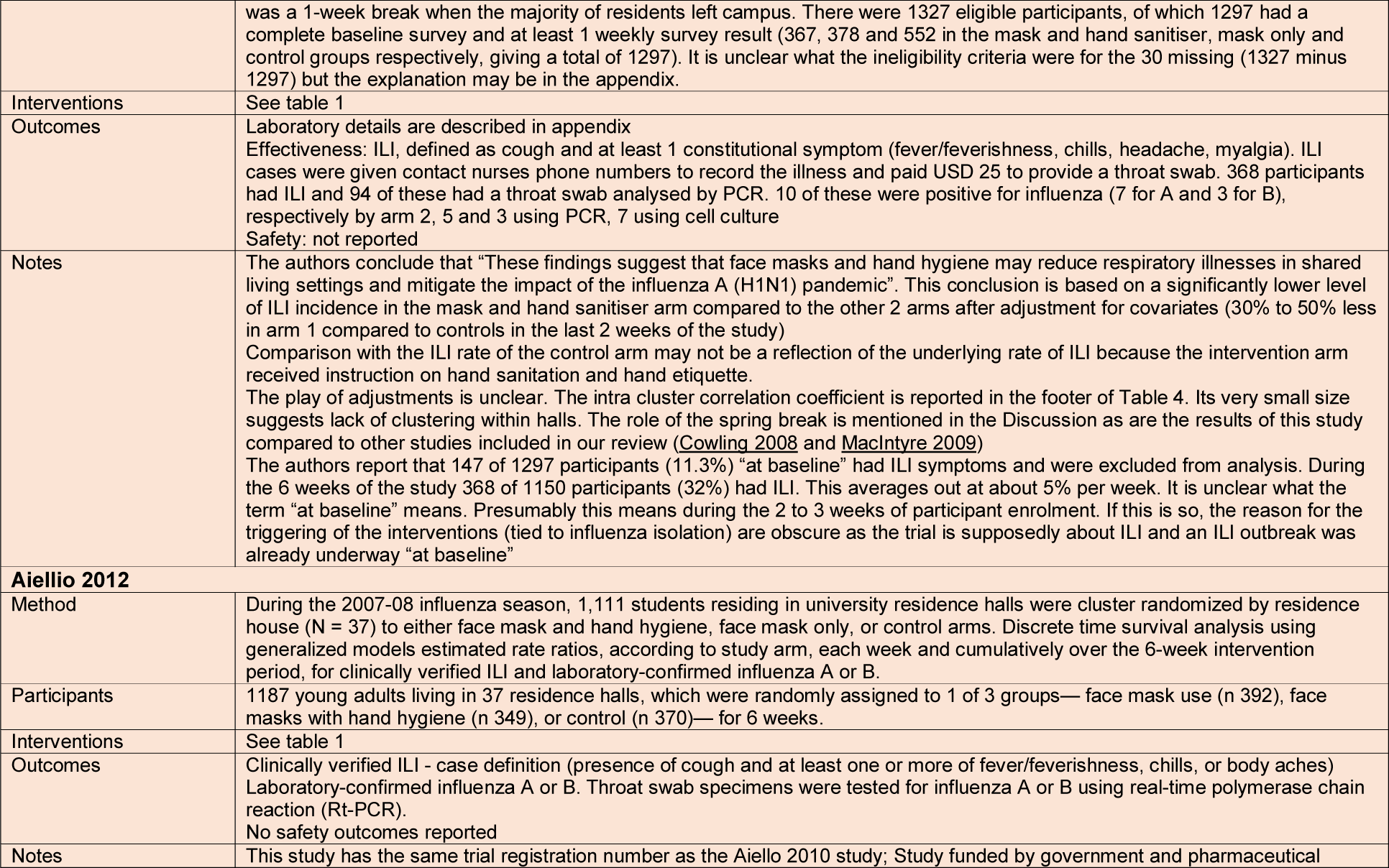
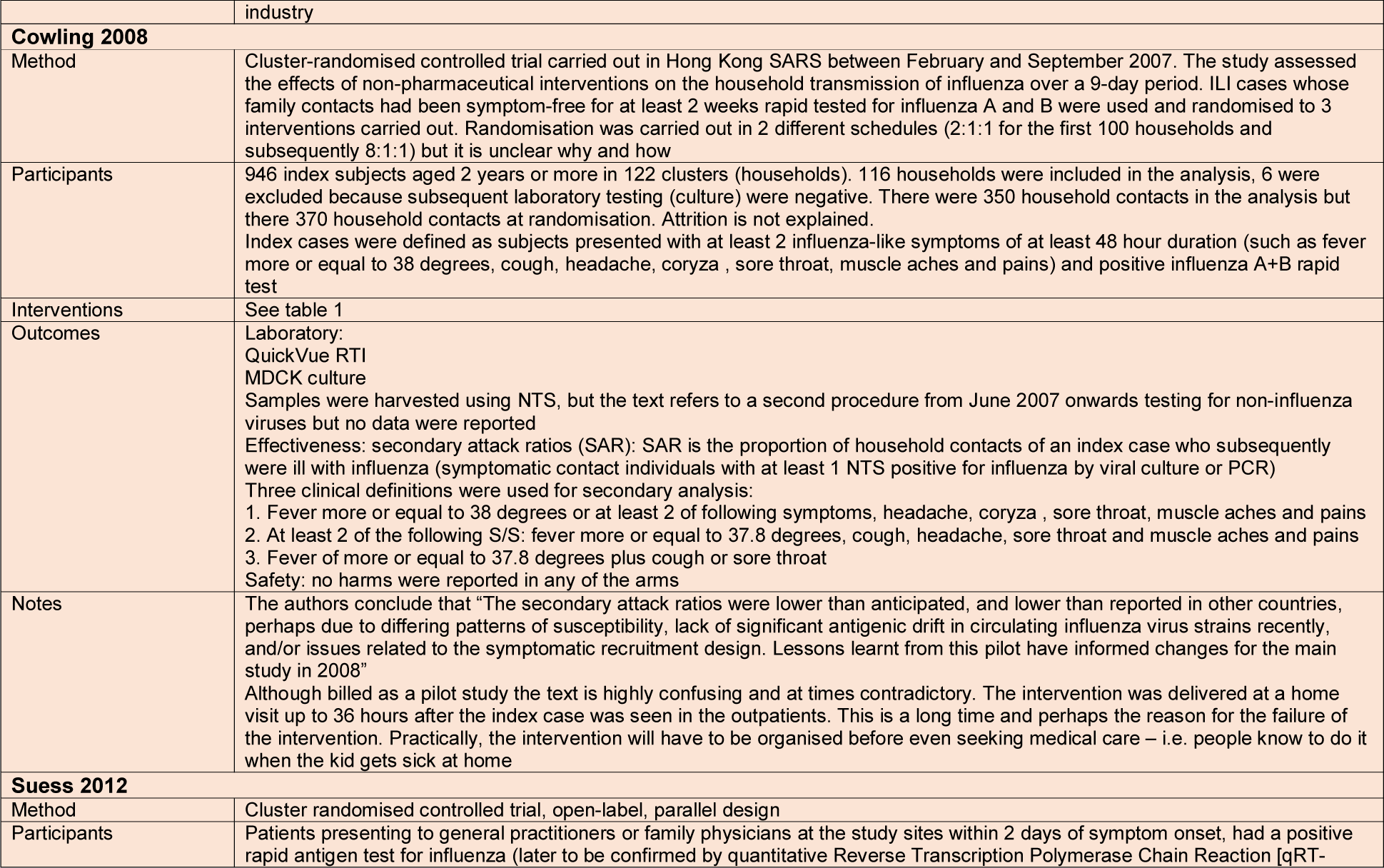
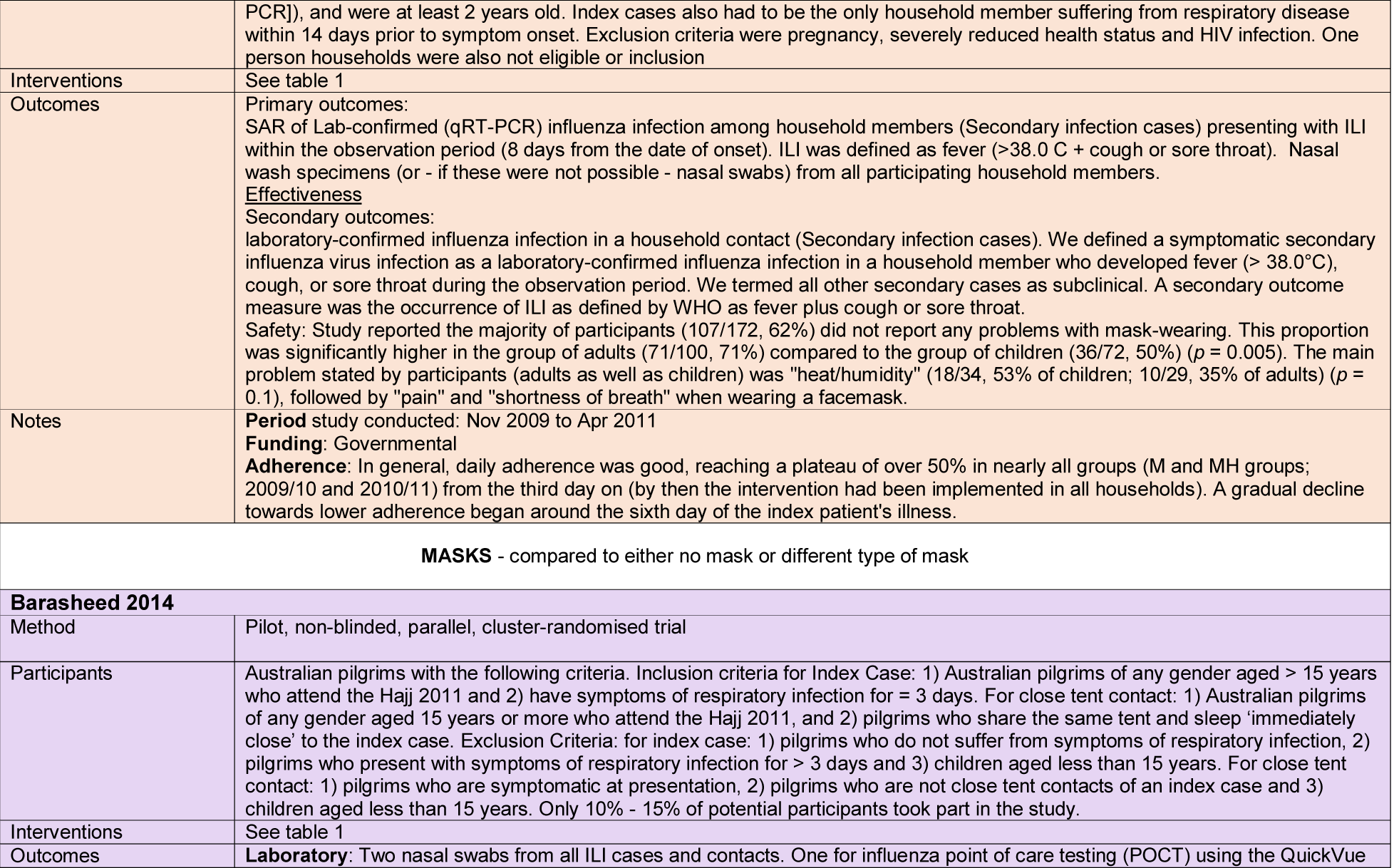
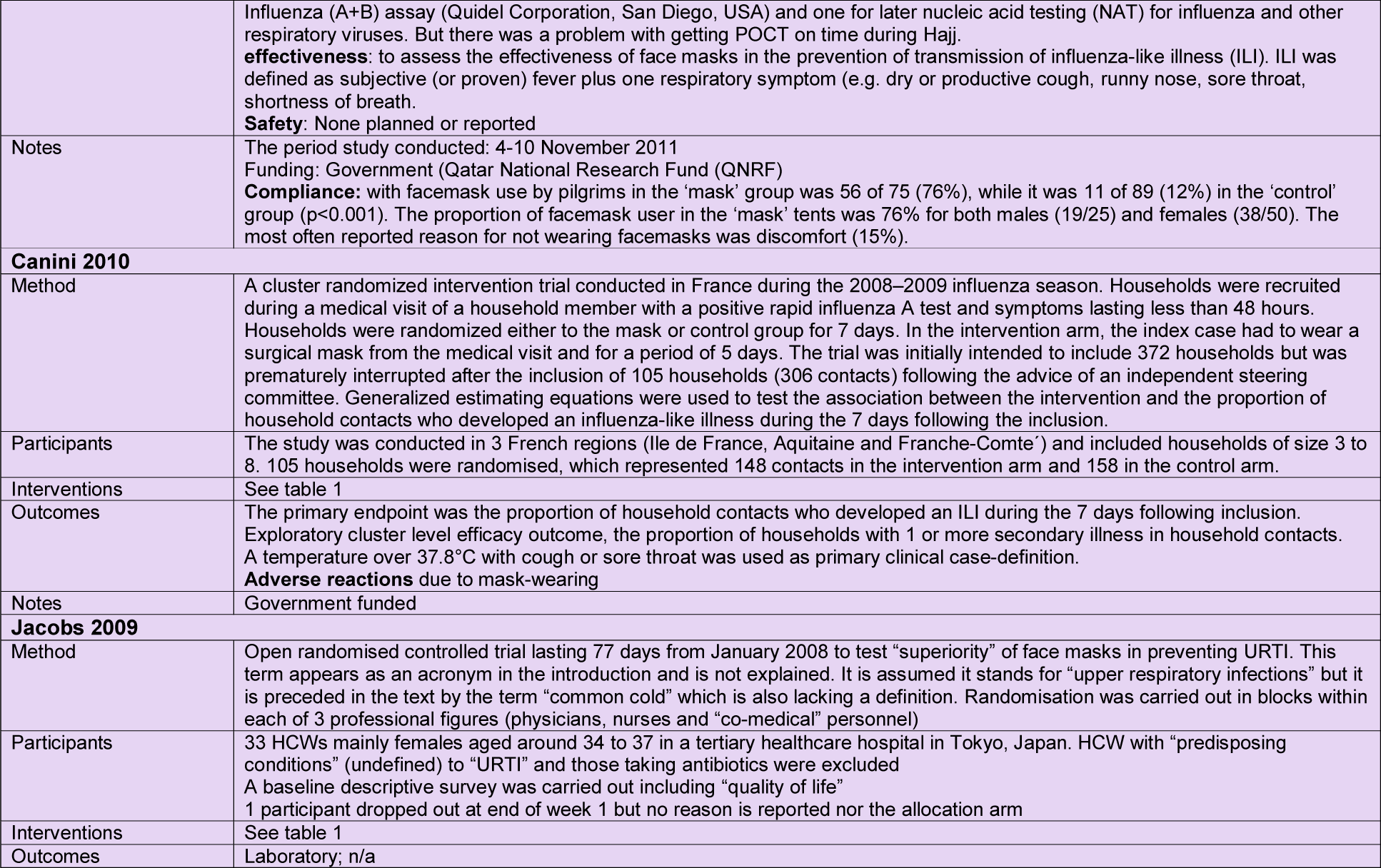
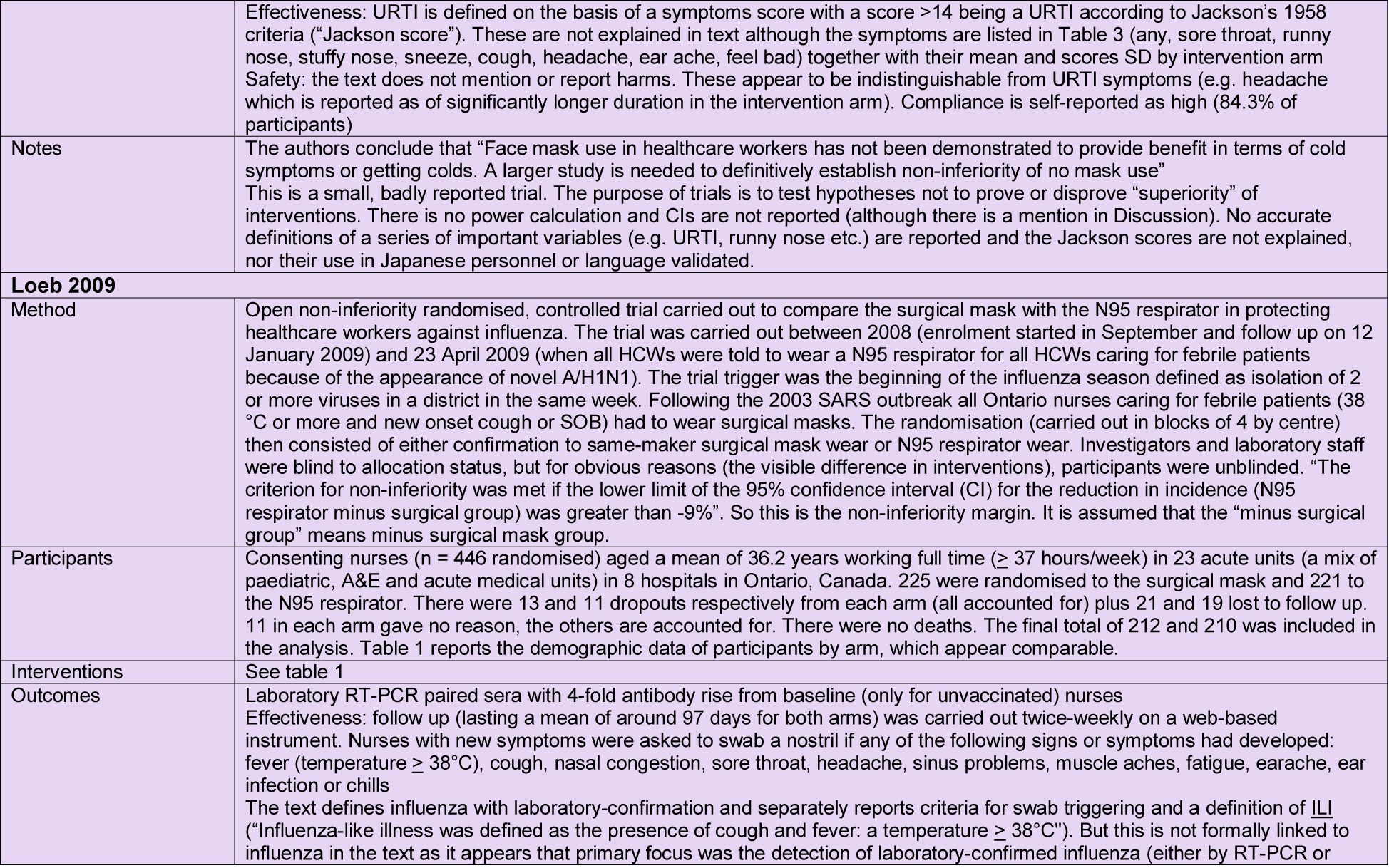
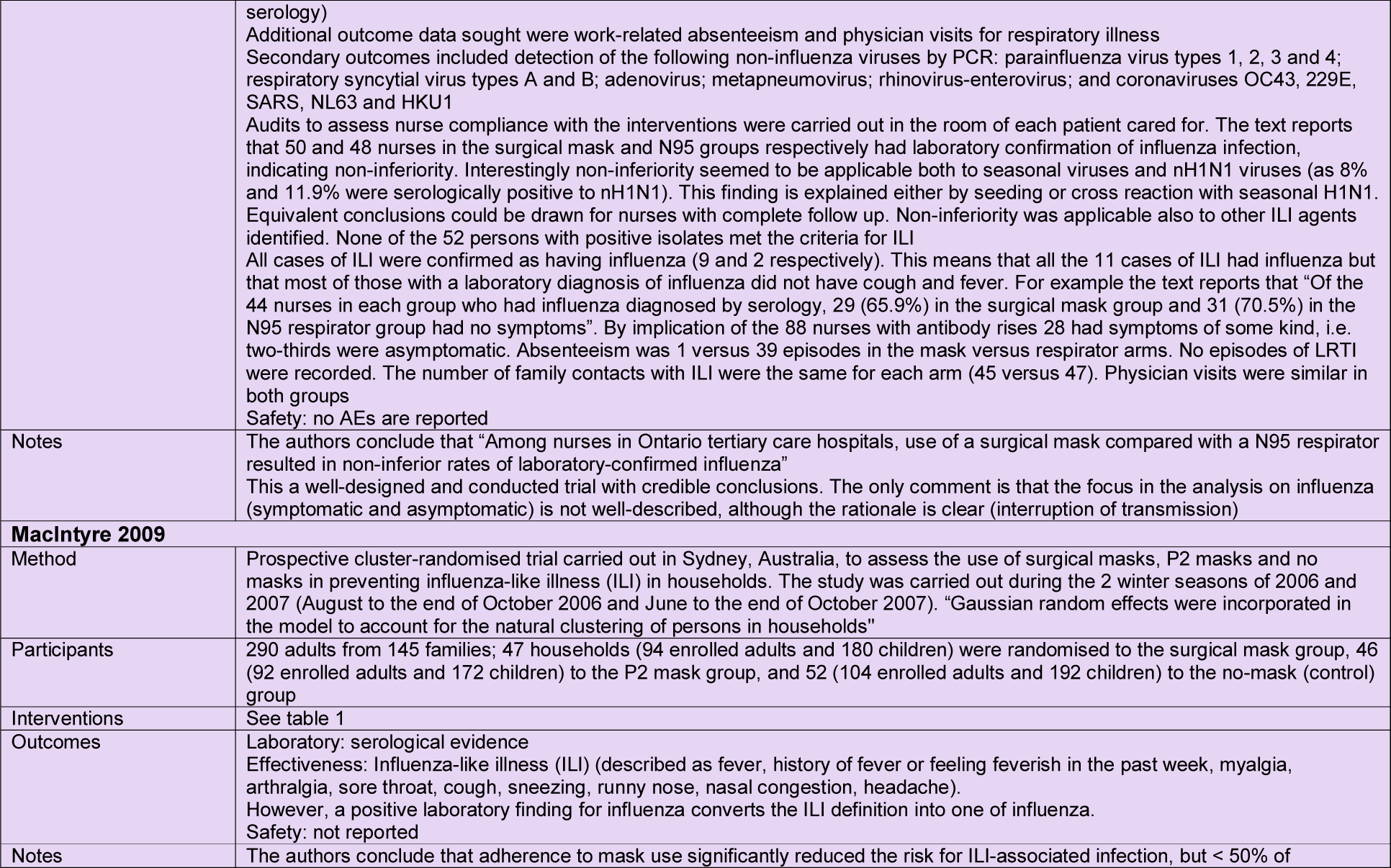
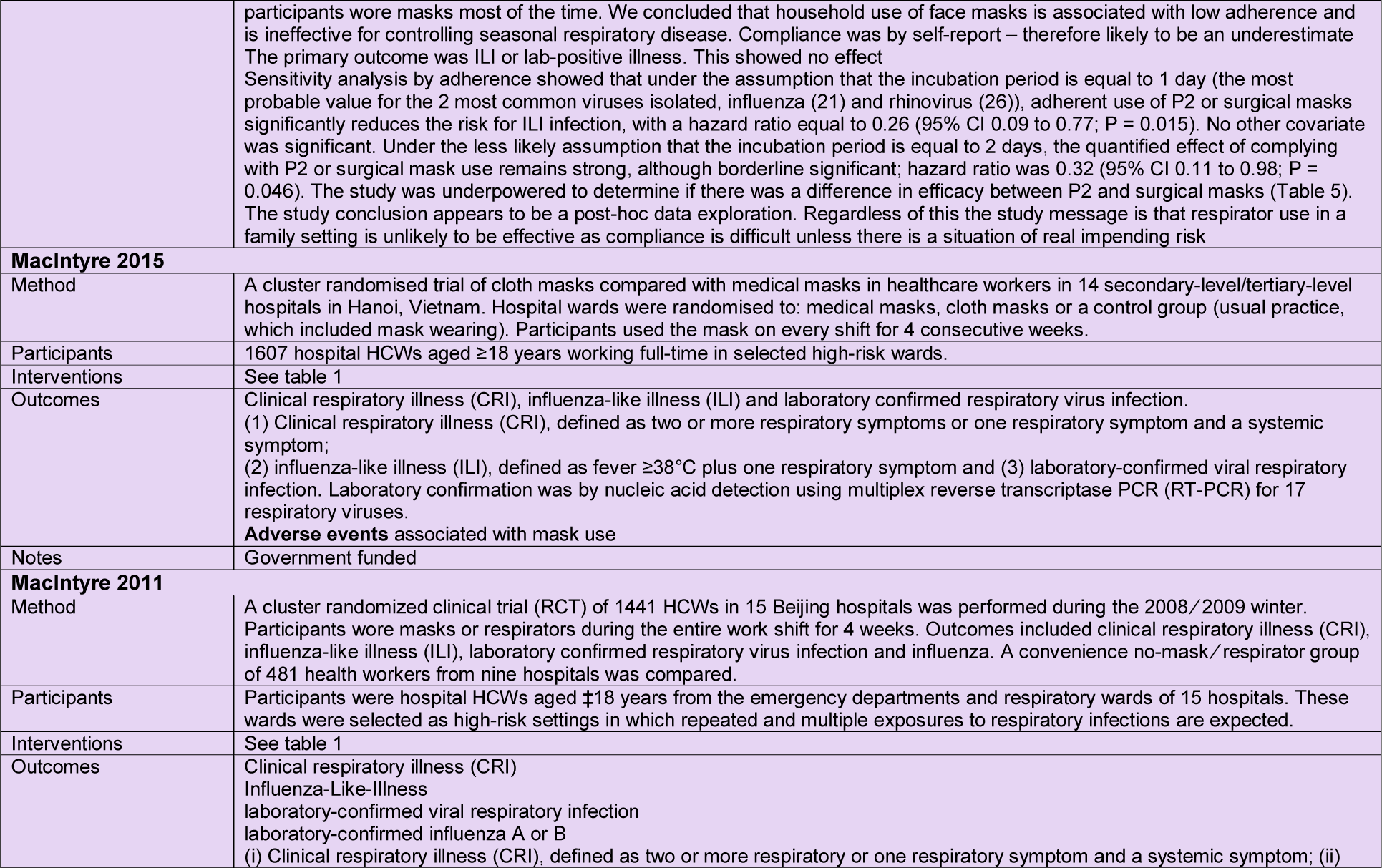
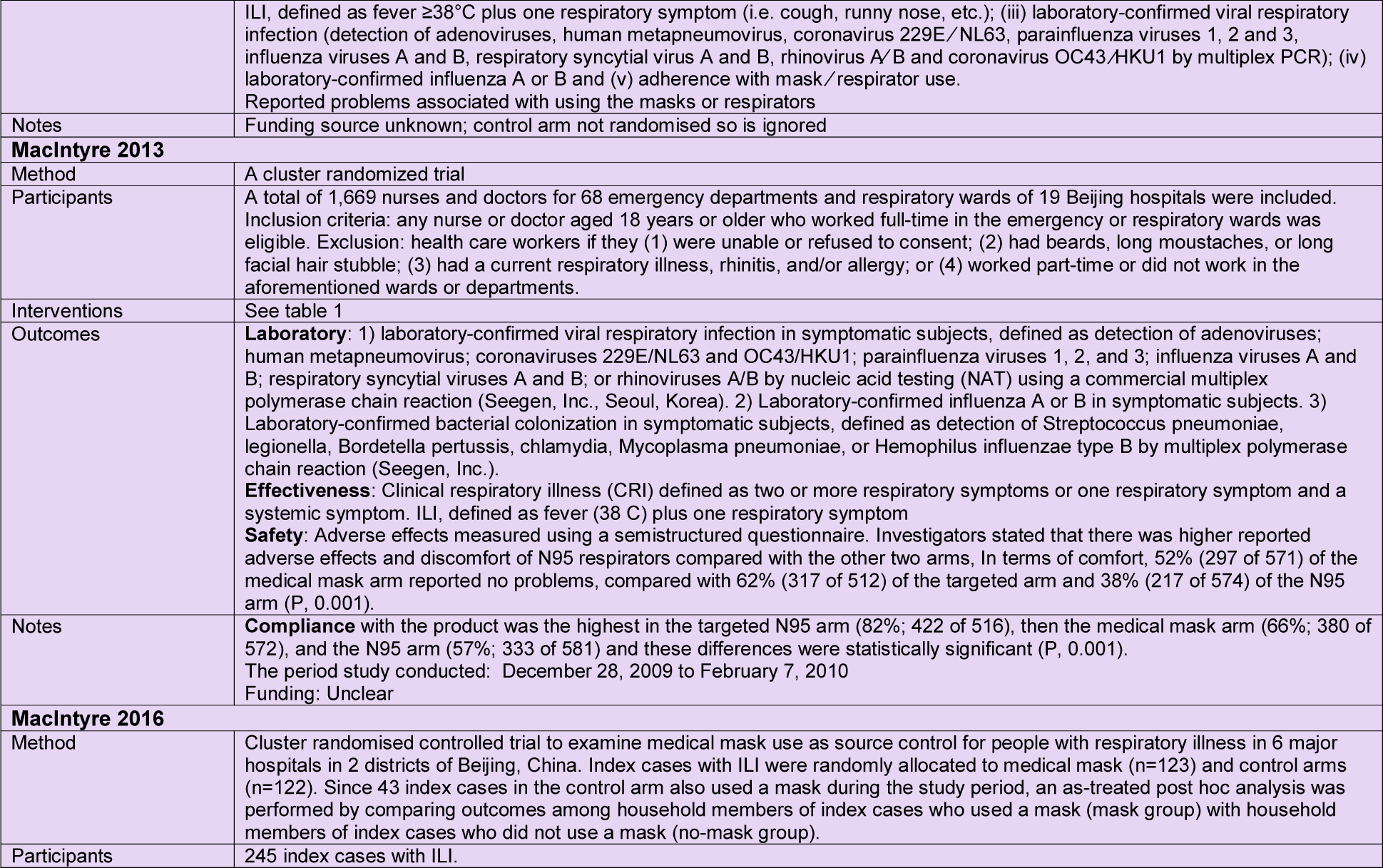
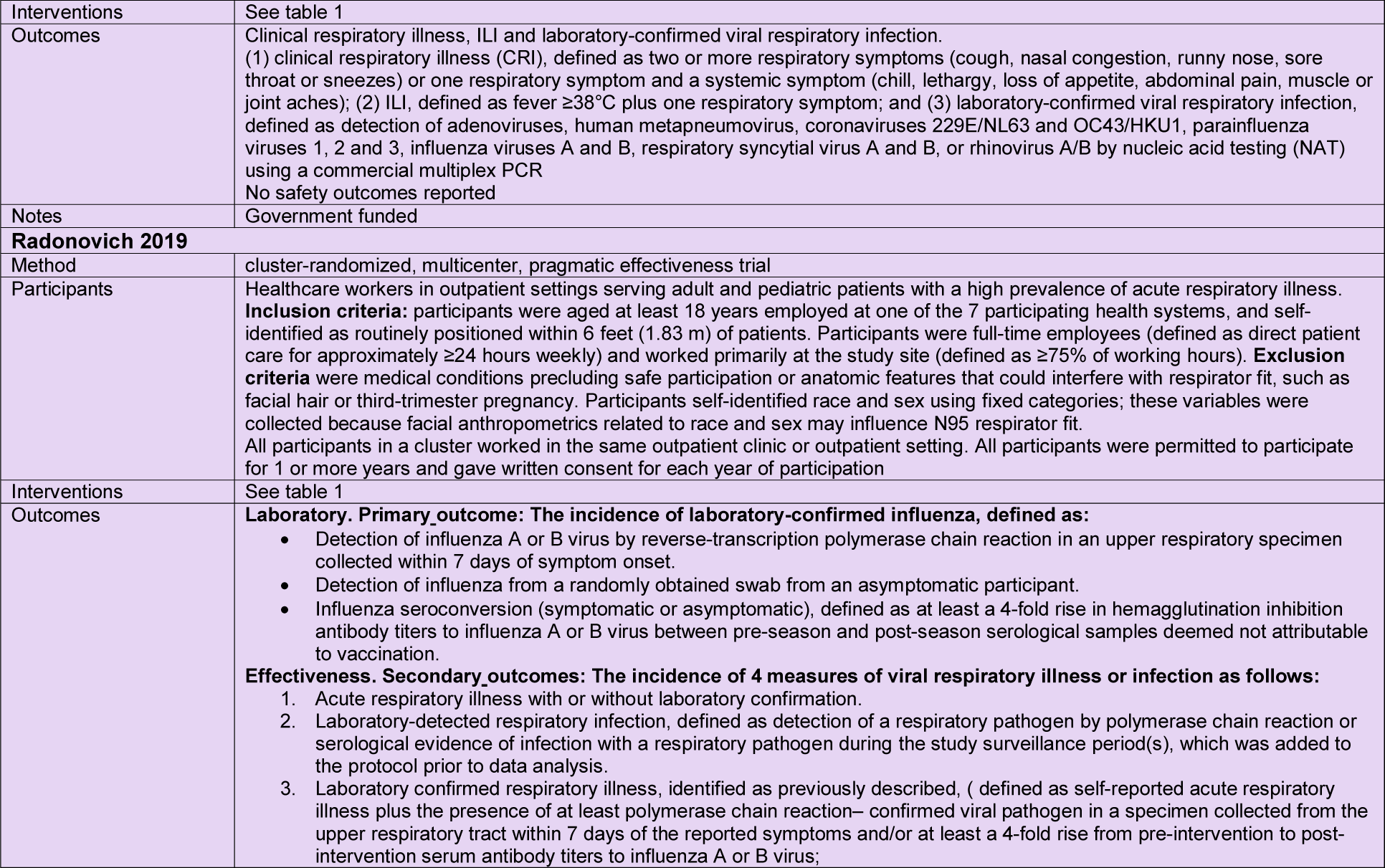
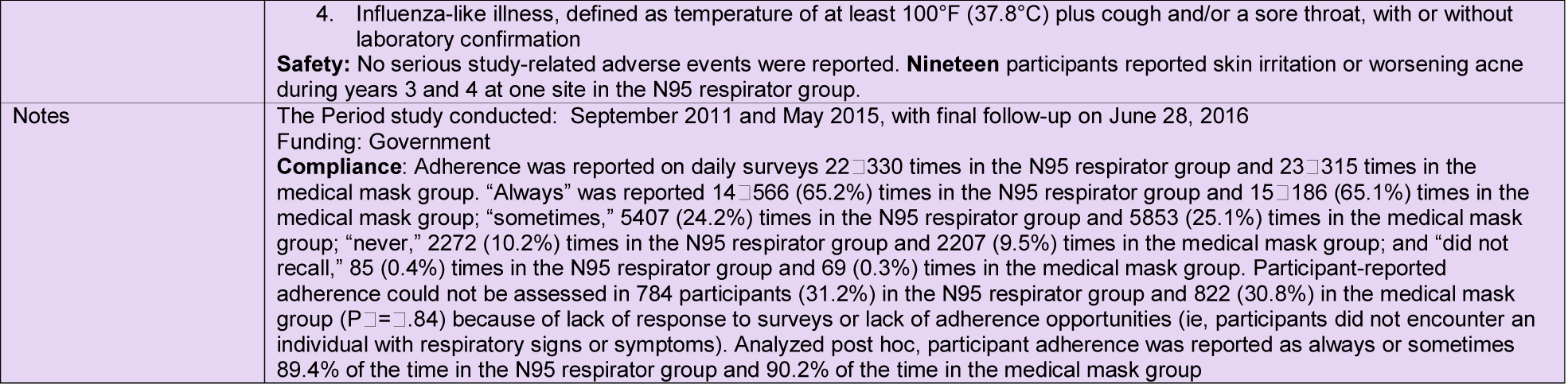
Characteristics of included studies

**Figure 3a:**
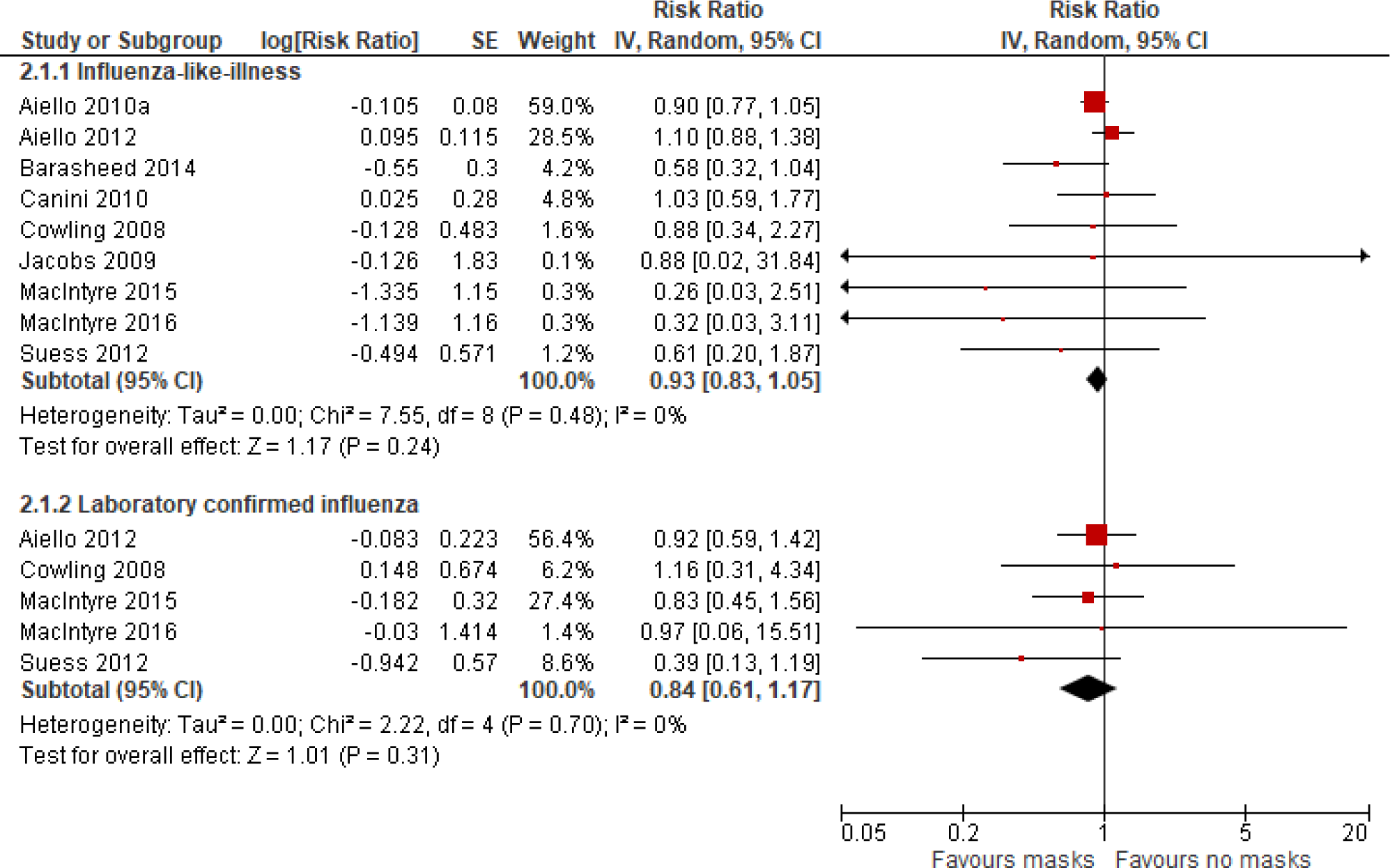
Meta-analysis of trials comparing masks vs no masks in healthcare workers and in community populations: effect on rates of Influenza-like Illness (ILI) and laboratory-confirmed influenza

Five trials compared surgical masks with N95/P2 respirators. ^25 27-30^ All trials except MacIntyre 2009^20^ included healthcare workers. Pooling of four trials showed no difference between surgical/medical face masks and N95 respirators for rates of clinical respiratory illness (Risk Ratio 0.70, 95%CI 0.45 to 1.10), for ILI (Risk Ratio 0.83, 95%CI 0.63 to 1.08), or for laboratory-confirmed influenza (Risk Ratio 1.02, 95%CI 0.73 to 1.43) (see Figure 3c). If only studies in healthcare workers are compared the Risk Ratio for ILI is 0.64, 95%CI 0.32 to 1.31. The outcomes ‘clinical respiratory illness’ and ILI were reported separately by the authors. Considering how these outcomes were defined it is highly likely that there is considerable overlap between the two and therefore these outcomes were not combined into a single clinical outcome. Harms were poorly reported, but generally discomfort wearing masks was mentioned^24^ and Radonovich^30^ mentioned that participants wearing the N95 respirator reported skin irritation and worsening of acne.

**Figure 3b:**
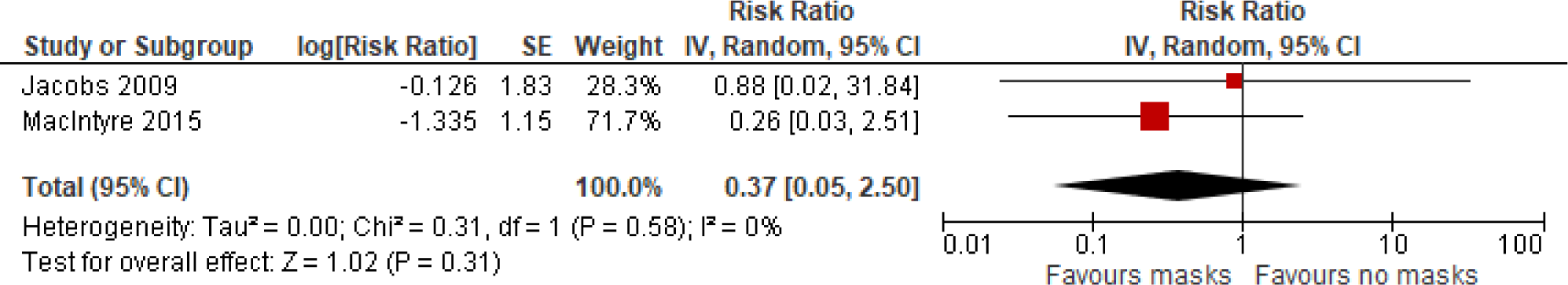
Meta-analysis of trials comparing masks vs no masks in healthcare workers: effect on rates of Influenza-like Illness (ILI)

**Figure 3c:**
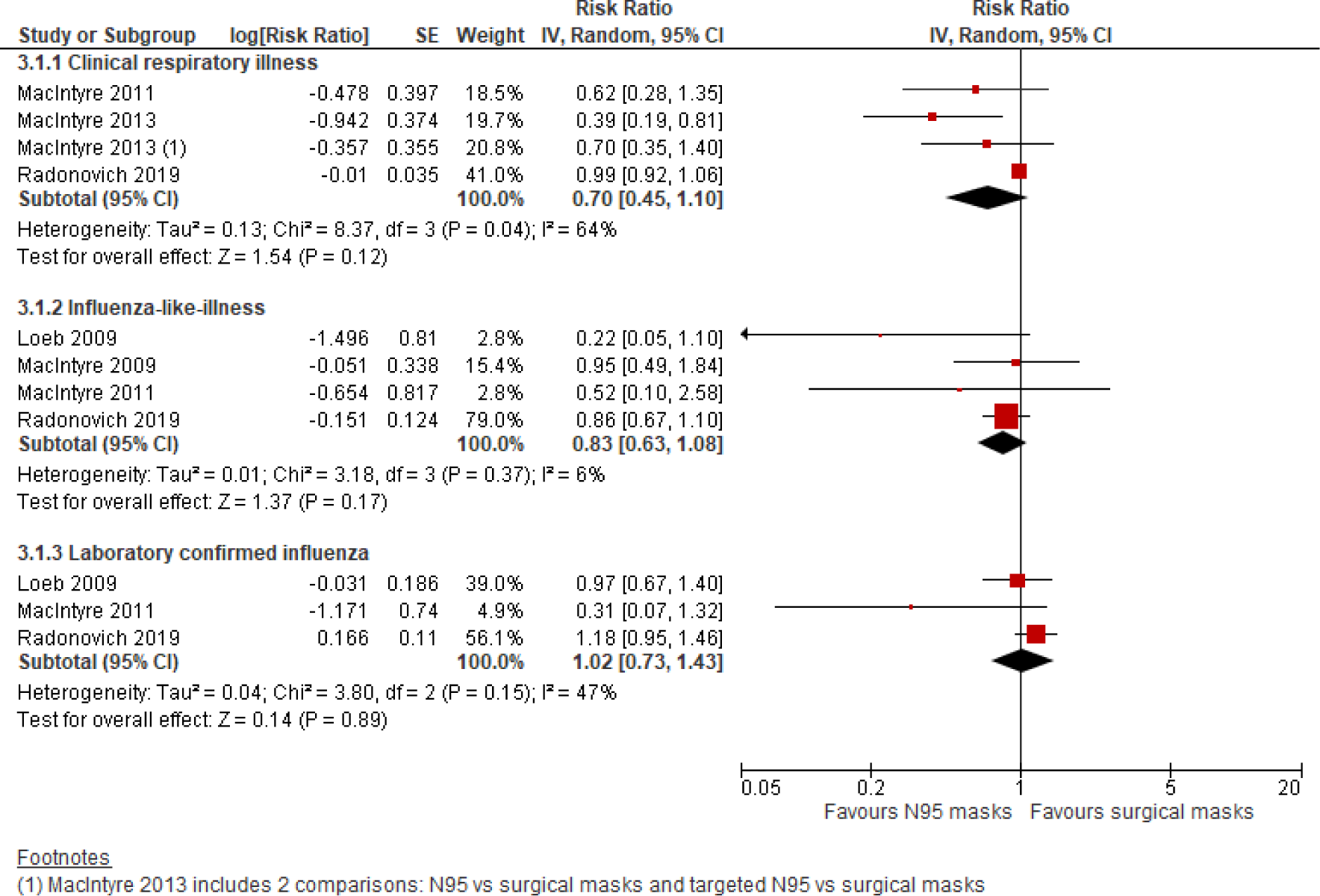
Meta-analysis of trials comparing (surgical/medical) masks vs N95 respirators in healthcare workers and in community populations: effect on rates of clinical respiratory illness, Influenza-like Illness (ILI) and laboratory-confirmed influenza

MacIntyre 2015^25^ also included a trial arm with cloth masks and found that the rate of ILI was higher in the cloth mask arm compared to medical/surgical masks (RR 13.25, 95%CI 1.74 to 100.97) and compared to no masks (RR 3.49, 95%CI 1.00 to 12.17).

### Person distancing

One trial evaluated the effectiveness of quarantining workers of one of two sibling companies in Japan whose family members developed an influenza-like Illness (ILI) during the 2009-2010 H1N1 influenza pandemic. ^31^ Workers in the intervention group were asked to stay home on full pay until 5 days after the household member(s) showed resolution of symptoms or 2 days after alleviation of fever. Compliance was 100%. In the intervention group 2.75% of workers contracted influenza, compared with 3.18% in the control group (Cox Hazard Ratio 0.799, 95%CI 0.658 to 0.970, p=0.023), indicating a 20% reduction of infection in the intervention group. However, the risk of a worker being infected was 2.17-fold higher in the intervention group where workers stayed at home with their infected family members. The authors conclude that quarantining workers with infected household members could be a useful additional measure to control spread of respiratory viruses in an epidemic setting.

## Discussion

### Main findings

Our results show that masks alone have no significant effect in interrupting spread of ILI RR 0.93 (95% CI 0.83 to 1.05) or influenza RR 0.84 (95% CI 0.61 to 1.17) in the all populations analysis. Our findings are similar for ILI in healthcare workers RR 0.37 (95% CIs 0.05 to 2.50) and for the comparisons between N95 respirators and surgical masks: RR 0.70 (95% CI 0.45 to 1.10) for clinical respiratory illness, RR 0.83 (95% CI 0.63 to 1.08) and influenza RR 1.02 (95% CI 0.73 to 1.43). Five of the trials contributing to the analysis were carried out by members of the same group. ^20 25 26 28 29^ On the basis of one trial^25^ cloth surgical masks should not be used as they are associated with a higher risk of ILI and penetration of microorganisms. In general, harms were not or poorly reported, with general discomfort resulting in reduced compliance with wearing being the main issues.

One trial testing person distancing found a reduction in transmission to co-workers when those with infected household members stay home from work. However, staying home increased their risk of being infected two-fold. We were disappointed to find only one trial on person distancing which is currently the core of the global containment strategy. This points to the difficulty and lack of interest in carrying out such studies.

Even though this update of the review focussed only on randomised and cluster randomised trials, the available body of evidence is inconclusive. We found a body of relatively small trials conducted mostly in a non-epidemic context (low viral circulation), with the exception of the largest study which crossed during the active study period two of the highest reporting years for influenza in the United States, between 2010-2017. ^32^ The two largest studies with respect to event rates are^19 30^ consistent regarding the direction of their findings of no differences between surgical or N95 masks.

Collectively, the evidence base was of variable quality. Inadequate reporting of sequence generation and allocation concealment was common. While allocation concealment of cluster-randomised trials is important this was rarely reported. Due to the nature of the intervention comparison, most trials were unblinded. However, blinding of outcome assessment is highly feasible and highly desirable, but was rarely done or reported. Outcomes were poorly defined with lack of clarity as to possible etiology of the agents (bacterial vs viral) in some studies. The cluster trials had insufficient attention paid to adjusting sample size calculations and analysis for clustering. As a consequence many trials were underpowered and had spuriously narrow confidence intervals around the effect size. The variable quality of the studies places some limits on the generalisability to the current COVID-19 epidemic, albeit it is a respiratory virus with a similar mode of transmission to those in the reviewed studies.

### Comparison with other reviews

In a meta-analysis comparing surgical masks with N95 respirators Smith^33^ pooled three trials^19 28 29^ and found no significant difference (OR 0.89, 95% CI 0.64 to 1.24) for laboratory-confirmed respiratory infections or ILI (OR 0.51, 95% CI 0.19 to 1.41). A similar meta-analysis by Offeddu et al^34^ concluded that based on two studies by MacIntyre et al^25 28^ masks (either surgical masks or N95 respirators) were effective against clinical respiratory infections (RR 0.59, 95% CI 0.46 to 0.77) and ILI (RR 0.34, 95% CI 0.14 to 0.82). Pooling the same two studies they also found that N95 respirators were superior to surgical masks for Clinical respiratory infections (RR 0.47, 95% CI 0.36 to 0.62), but not for ILI (RR 0.59, 95%CI 0.27 to 1.28). ^34^ The most recent meta-analysis by Long et al^35^ included 5 studies comparing surgical masks with N95 respirators and found no difference (RR 1.09, 95% CI 0.92 to 1.28) against neither influenza nor respiratory viral infections (RR 0.89, 95% CI 0.70 to 1.11).35 By excluding the Loeb^19^ study (an open non-inferiority randomised, controlled trial carried out to compare the surgical mask with the N95 respirator in protecting healthcare workers against influenza) the authors found a significant effect against viral infections (RR 0.61, 95% CI 0.39 to 0.98). The authors do not report a rationale for the exclusion in the sensitivity analysis and do not report on exclusion of the studies with low weighting which arguably would be more relevant in a sensitivity analysis. The two studies which make up 96% of the weighting^19 30^ clearly demonstrate no difference in the outcome events.

The findings from several systematic reviews and meta-analyses over the last decade have not demonstrated any significant difference in the clinical effectiveness of N95 respirators or equivalent compared to the use of surgical masks when used by healthcare workers in multiple health care settings for the prevention of respiratory virus infections, including influenza.

Our 2011 review^8^ showed a clear protective effect of wearing surgical masks and hygienic measures compared to not wearing masks in the SARS 2003 outbreak (RR 0.32, 95% CIs 0.26 to 0.39). The evidence was based on case-control studies carried out during the outbreak.

### Relevance of the Findings in the Clinical Setting

Our findings are highly relevant in the setting of epidemic and pandemic respiratory infections and the current global pandemic of COVID-19 underscores the point. The evidence supports that SARS-CoV-2 is spread through respiratory droplets and/or contact routes which places it in the route of transmission for which health care workers would be required to wear masks.^36 37^ The WHO China Joint Mission on COVID-19 of 75,465 cases supports person-to-person droplet and fomite transmission, with the majority of transmission occurring within families in close contact with each other. ^36^ A recent report in a clinical setting of intubation and non-invasive ventilation in which 41 health care workers were exposed over a prolonged period a within close proximity to a COVID-19 + patient revealed no transmission events to the Health care workers based on repeated testing during which majority (85%) of the health care workers were wearing a surgical mask and other appropriate PPE while the remainder wore an N95 respirator. This latter finding supports the results of the studies which were reviewed and brings into focus the importance of the use of masks as a component of personal protective equipment in the current COVID-19 pandemic. The current COVID-19 pandemic has elicited conflicting recommendations with several institutions and countries suggesting that only N95 or equivalent masks should be used as a component of the personal protective equipment for health care workers and not a surgical mask. The WHO recommendations emphasize that in the setting of epidemic and pandemic respiratory virus infections transmitted predominantly by the droplet route, one of the most important elements is strict adherence in the use of personal protective equipment of which the facial mask is only one component^38^ and suggest surgical masks for routine care and reserve the N95 mask for aerosol generating medical procedures. Despite the methodological issues outlined, our review of the available literature did not find any differences in the clinical effectiveness of either type of mask in the setting of respiratory viral infection transmission to health care workers. Our review also identified a dearth of reported findings related to the harms of N95 respirators and they need to be considered in any RCTs or C-RTs, especially in the setting of a global pandemic with the potential of frequent and prolonged use. Many such harms were identified in the setting of the SARS epidemic in 2003 and in the ensuing years and included respiratory fatigue, increased work of breathing, poor work capability, increased nasal resistance, fatigue with minimal workloads, elevated levels of carbon dioxide, facial dermatitis, acne and potential self-contamination events.^39-44^

### Limitations

Though the trials in this review provide a reasonable body of evidence, there are several important limitations. First, there is considerable clinical heterogeneity between the designs, and substantial statistical heterogeneity for some analyses. The latter is not readily explained by differences in the study questions. Second, the range of viral infections studied is limited, with a particular focus on influenza; no studies include SARS-CoV-2. None of the studies in health care worker included undertaking aerosol-generating procedures for which WHO currently recommends the N95 or equivalent mask. Finally, the studies provide sparse and unsystematic data on any harms, such as the discomfort, dehydration, facial dermatitis, distress, headaches, exhaustion or other problems caused by masks such as the N95. Some studies measured adherence which was generally high despite the mask discomfort.

### Conclusion

Despite the lack of evidence, we would still recommend using facial barriers in the setting of epidemic and pandemic viral respiratory infections, but there does not appear to be a difference between surgical and full respirator wear. Despite the methodological concerns, our review of the available studies demonstrates consistency in the finding of no difference between surgical and N95 or equivalent masks as a physical intervention to interrupt or reduce the spread of respiratory viruses, mainly influenza. The consistency of the finding across multiple studies of variable quality adds epidemiologic strength of association.

The fact that all included trials were conducted in relatively low transmission periods limits generalisability to an epidemic of the global size of COVID-19. We excluded in this part of the review trials testing the combination of hygienic and barrier methods. These have shown to be effective in observational studies carried out during the SARS 1 epidemic.^8^

Based on the evidence of the previous SARS epidemic large trials comparing full facial protection with surgical masks need to be carried out to settle the matter, given the difference in wearability, harms and costs. Funding for such trials and research once the epidemic has passed, is critical to inform future preparedness for global epidemics.

## Data Availability

All data in this review are from published journal articles. Extraction sheets are available from correspoding author.

https://www.cochranelibrary.com/cdsr/doi/10.1002/14651858.CD006207.pub4/full

## Disclosure

Tom Jefferson (TJ) was in receipt of a Cochrane Methods Innovations Fund grant to develop guidance on the use of regulatory data in Cochrane reviews (2015-018).

In 2014–2016, TJ was a member of three advisory boards for Boehringer Ingelheim. TJ was a member of an independent data monitoring committee for a Sanofi Pasteur clinical trial on an influenza vaccine.

TJ is occasionally interviewed by market research companies about phase I or II pharmaceutical products for which he receives fees (current).

TJ was a member of three advisory boards for Boehringer Ingelheim (2014-16)

TJ was a member of an independent data monitoring committee for a Sanofi Pasteur clinical trial on an influenza vaccine (2015-2017).

TJ is a relator in a False Claims Act lawsuit on behalf of the United States that involves sales of Tamiflu for pandemic stockpiling. If resolved in the United States’ favor, he would be entitled to a percentage of the recovery.

TJ is co-holder of a Laura and John Arnold Foundation grant for development of a RIAT support centre (2017-2020) and Jean Monnet Network Grant, 2017-2020 for The Jean Monnet Health Law and Policy Network. TJ is an unpaid collaborator to the project *Beyond Transparency in Pharmaceutical Research and Regulation* led by Dalhousie University and funded by the Canadian Institutes of Health Research (2018-2022).

TJ consults for Illumina LLC on next generation gene sequencing (2019-). TJ was the consultant scientific coordinator for the HTA Medical Technology programme of the Agenzia per i Servizi Sanitari Nazionali (AGENAS) of the Italian MoH (2007-2019).

TJ is Director Medical Affairs for BC Solutions, a market access company for medical devices in Europe (excluding devices relating to acute respiratory infections).

John Conly holds grants from the Canadian Institutes for Health Research, Alberta Innovates-Health Solutions and was the primary local Investigator for a *Staphylococcus aureus* vaccine study funded by Pfizer for which all funding was provided only to the University of Calgary for the conduct of the trial.

All other authors have no interests to declare.

## Contributorship

All authors contributed equally to the design of the update, screening, extraction, interpretation and writing the manuscript which as approved by all authors. JC designed and carried out the searches and MA and EB carried out the analysis.

## Acknowledgements

The authors thank Dr Elizabeth Gibson for her assistance with data extraction.

The sponsors had no role in any aspect of preparation of the manuscript.

Dr Jefferson (the manuscript’s guarantor) affirms that the manuscript is an honest, accurate, and transparent account of the study being reported; that no important aspects of the study have been omitted; and that any discrepancies from the study as planned (and, if relevant, registered) have been explained.

## Appendix

### Appendix 1: Search strings for databases

#### PubMed search run 09/03/2020

(“Influenza, Human”[Mesh] OR “Influenzavirus A”[Mesh] OR “Influenzavirus B”[Mesh] OR “Influenzavirus C”[Mesh] OR Influenza[tiab] OR “Respiratory Tract Diseases”[Mesh] OR “Bacterial Infections/transmission”[Mesh] OR Influenzas[tiab] OR “Influenza-like”[tiab] OR ILI[tiab] OR Flu[tiab] OR Flus[tiab] OR “Common Cold”[Mesh:NoExp] OR “common cold”[tiab] OR colds[tiab] OR coryza[tiab] OR coronavirus[Mesh] OR “sars virus”[Mesh] OR coronavirus[tiab] OR Coronaviruses[tiab] OR “coronavirus infections”[Mesh] OR “severe acute respiratory syndrome”[Mesh] OR “severe acute respiratory syndrome”[tiab] OR “severe acute respiratory syndromes”[tiab] OR sars[tiab] OR “respiratory syncytial viruses”[Mesh] OR “respiratory syncytial virus, human”[Mesh] OR “Respiratory Syncytial Virus Infections”[Mesh] OR “respiratory syncytial virus”[tiab] OR “respiratory syncytial viruses”[tiab] OR rsv[tiab] OR parainfluenza[tiab] OR ((Transmission[tiab]) AND (Coughing[tiab] OR Sneezing[tiab])) OR ((respiratory[tiab] AND Tract[tiab]) AND (infection[tiab] OR Infections[tiab] OR illness[tiab])))

AND

(“Hand Hygiene”[Mesh] OR handwashing[tiab] OR hand-washing[tiab] OR ((Hand[tiab] OR Alcohol[tiab]) AND (wash[tiab] OR Washing[tiab] OR Cleansing[tiab] OR Rinses[tiab] OR hygiene[tiab] OR rub[tiab] OR Rubbing[tiab] OR sanitiser[tiab] OR sanitizer[tiab] OR cleanser[tiab] OR disinfected[tiab] OR Disinfectant[tiab] OR Disinfect[tiab] OR antiseptic[tiab] OR virucid[tiab])) OR “gloves, protective”[Mesh] OR Glove[tiab] OR Gloves[tiab] OR Masks[Mesh] OR “respiratory protective devices”[Mesh] OR facemask[tiab] OR Facemasks[tiab] OR mask[tiab] OR Masks[tiab] OR respirator[tiab] OR respirators[tiab] OR “Protective Clothing”[Mesh:NoExp] OR “Protective Devices”[Mesh] OR “patient isolation”[tiab] OR ((school[tiab] OR Schools[tiab]) AND (Closure[tiab] OR Closures[tiab] OR Closed[tiab])) OR Quarantine[Mesh] OR quarantine[tiab] OR “Hygiene intervention”[tiab] OR “Mouthwashes”[Mesh] OR gargling[tiab] OR “nasal tissues”[tiab])

AND

(“Communicable Disease Control”[Mesh] OR “Disease Outbreaks”[Mesh] OR “Disease Transmission, Infectious”[Mesh] OR “Infection Control”[Mesh] OR Transmission[sh] OR “Prevention and control”[sh] OR “Communicable Disease Control”[tiab] OR “Secondary transmission”[tiab] OR ((Reduced[tiab] OR Reduce[tiab] OR Reduction[tiab] OR Reducing[tiab] OR Lower[tiab]) AND (Incidence[tiab] OR Occurrence[tiab] OR Transmission[tiab] OR Secondary[tiab])))

AND

(Randomized controlled trial[pt] OR controlled clinical trial[pt] OR randomized[tiab] OR randomised[tiab] OR placebo[tiab] OR “drug therapy”[sh] OR randomly[tiab] OR trial[tiab] OR groups[tiab])

NOT

(Animals[Mesh] not (Animals[Mesh] and Humans[Mesh]))

NOT

(“Case Reports”[pt] OR Editorial[pt] OR Letter[pt] OR Meta-Analysis[pt] OR “Observational Study”[pt] OR “Systematic Review”[pt] OR “Case Report”[ti] OR “Case series”[ti] OR Meta-Analysis[ti] OR “Meta Analysis”[ti] OR “Systematic Review”[ti])

#### Cochrane CENTRAL run 09/03/2020

([mh “Influenza, Human”] OR [mh “Influenzavirus A”] OR [mh “Influenzavirus B”] OR [mh “Influenzavirus C”] OR Influenza:ti,ab OR [mh “Respiratory Tract Diseases”] OR Influenzas:ti,ab OR “Influenza-like”:ti,ab OR ILI:ti,ab OR Flu:ti,ab OR Flus:ti,ab OR [mh ^”Common Cold”] OR “common cold”:ti,ab OR colds:ti,ab OR coryza:ti,ab OR [mh coronavirus] OR [mh “sars virus”] OR coronavirus:ti,ab OR Coronaviruses:ti,ab OR [mh “coronavirus infections”] OR [mh “severe acute respiratory syndrome”] OR “severe acute respiratory syndrome”:ti,ab OR “severe acute respiratory syndromes”:ti,ab OR sars:ti,ab OR [mh “respiratory syncytial viruses”] OR [mh “respiratory syncytial virus, human”] OR [mh “Respiratory Syncytial Virus Infections”] OR “respiratory syncytial virus”:ti,ab OR “respiratory syncytial viruses”:ti,ab OR rsv:ti,ab OR parainfluenza:ti,ab OR ((Transmission) AND (Coughing OR Sneezing)) OR ((respiratory:ti,ab AND Tract) AND (infection:ti,ab OR Infections:ti,ab OR illness:ti,ab)))

AND

([mh “Hand Hygiene”] OR handwashing:ti,ab OR “hand-washing”:ti,ab OR ((Hand:ti,ab OR Alcohol:ti,ab) AND (wash:ti,ab OR Washing:ti,ab OR Cleansing:ti,ab OR Rinses:ti,ab OR hygiene:ti,ab OR rub:ti,ab OR Rubbing:ti,ab OR sanitiser:ti,ab OR sanitizer:ti,ab OR cleanser:ti,ab OR disinfected:ti,ab OR Disinfectant:ti,ab OR Disinfect:ti,ab OR antiseptic:ti,ab OR virucid:ti,ab)) OR [mh “gloves, protective”] OR Glove:ti,ab OR Gloves:ti,ab OR [mh Masks] OR [mh “respiratory protective devices”] OR facemask:ti,ab OR Facemasks:ti,ab OR mask:ti,ab OR Masks:ti,ab OR respirator:ti,ab OR respirators:ti,ab OR [mh ^”Protective Clothing”] OR [mh “Protective Devices”] OR “patient isolation”:ti,ab OR ((school:ti,ab OR Schools:ti,ab) AND (Closure:ti,ab OR Closures:ti,ab OR Closed:ti,ab)) OR [mh Quarantine] OR quarantine:ti,ab OR “Hygiene intervention”:ti,ab OR [mh Mouthwashes] OR gargling:ti,ab OR “nasal tissues”:ti,ab)

AND

([mh “Communicable Disease Control”] OR [mh “Disease Outbreaks”] OR [mh “Disease Transmission, Infectious”] OR [mh “Infection Control”] OR “Communicable Disease Control”:ti,ab OR “Secondary transmission”:ti,ab OR ((Reduced:ti,ab OR Reduce:ti,ab OR Reduction:ti,ab OR Reducing:ti,ab OR Lower:ti,ab) AND (Incidence:ti,ab OR Occurrence:ti,ab OR Transmission:ti,ab OR Secondary:ti,ab)))

#### Embase run 09/03/2020

(‘influenza’/exp OR Influenza:ti,ab OR ‘Respiratory Tract Disease’/exp OR Influenzas:ti,ab OR Influenza-like:ti,ab OR ILI:ti,ab OR Flu:ti,ab OR Flus:ti,ab OR ‘Common Cold’/de OR “common cold”:ti,ab OR colds:ti,ab OR coryza:ti,ab OR ‘coronavirus’/exp OR ‘SARS coronavirus’/exp OR coronavirus:ti,ab OR Coronaviruses:ti,ab OR ‘coronavirus infection’/exp OR ‘severe acute respiratory syndrome’/exp OR “severe acute respiratory syndrome”:ti,ab OR “severe acute respiratory syndromes”:ti,ab OR sars:ti,ab OR ‘Pneumovirus’/exp OR ‘Human respiratory syncytial virus’/exp OR “respiratory syncytial virus”:ti,ab OR “respiratory syncytial viruses”:ti,ab OR rsv:ti,ab OR parainfluenza:ti,ab OR ((Transmission) AND (Coughing OR Sneezing)) OR ((respiratory:ti,ab AND Tract) AND (infection:ti,ab OR Infections:ti,ab OR illness:ti,ab)))

AND

(‘hand washing’/exp OR handwashing:ti,ab OR hand-washing:ti,ab OR ((Hand:ti,ab OR Alcohol:ti,ab) AND (wash:ti,ab OR Washing:ti,ab OR Cleansing:ti,ab OR Rinses:ti,ab OR hygiene:ti,ab OR rub:ti,ab OR Rubbing:ti,ab OR sanitiser:ti,ab OR sanitizer:ti,ab OR cleanser:ti,ab OR disinfected:ti,ab OR Disinfectant:ti,ab OR Disinfect:ti,ab OR antiseptic:ti,ab OR virucid:ti,ab)) OR ‘protective glove’/exp OR Glove:ti,ab OR Gloves:ti,ab OR ‘mask’/exp OR ‘gas mask’/exp OR facemask:ti,ab OR Facemasks:ti,ab OR mask:ti,ab OR Masks:ti,ab OR respirator:ti,ab OR respirators:ti,ab OR ‘protective clothing’/de OR ‘protective equipment’/exp OR “patient isolation”:ti,ab OR ((school:ti,ab OR Schools:ti,ab) AND (Closure:ti,ab OR Closures:ti,ab OR Closed:ti,ab)) OR ‘Quarantine’/exp OR quarantine:ti,ab OR “Hygiene intervention”:ti,ab OR ‘mouthwash’/exp OR gargling:ti,ab OR “nasal tissues”:ti,ab)

AND

(‘Communicable Disease Control’/exp OR ‘epidemic’/exp OR ‘disease transmission’/exp OR ‘Infection Control’/exp OR “Communicable Disease Control”:ti,ab OR “Secondary transmission”:ti,ab OR ((Reduced:ti,ab OR Reduce:ti,ab OR Reduction:ti,ab OR Reducing:ti,ab OR Lower:ti,ab) AND (Incidence:ti,ab OR Occurrence:ti,ab OR Transmission:ti,ab OR Secondary:ti,ab)))

AND

(random* OR factorial OR crossover OR placebo OR blind OR blinded OR assign OR assigned OR allocate OR allocated OR ‘crossover procedure’/exp OR ‘double-blind procedure’/exp OR ‘randomized controlled trial’/exp OR ‘single-blind procedure’/exp NOT (‘animal’/exp NOT (‘animal’/exp AND ‘human’/exp)))

#### CINAHL run 09/03/2020

((MH “Influenza, Human+”) OR (MH “Orthomyxoviridae+”) OR TI Influenza OR AB Influenza OR (MH “Respiratory Tract Diseases+”) OR TI Influenzas OR AB Influenzas OR TI Influenza-like OR AB Influenza-like OR TI ILI OR AB ILI OR TI Flu OR AB Flu OR TI Flus OR AB Flus OR (MH “Common Cold+”) OR TI “common cold” OR AB “common cold” OR TI colds OR AB colds OR TI coryza OR AB coryza OR (MH “coronavirus+”) OR (MH “sars virus+”) OR TI coronavirus OR AB coronavirus OR TI Coronaviruses OR AB Coronaviruses OR (MH “coronavirus infections+”) OR (MH “severe acute respiratory syndrome+”) OR TI “severe acute respiratory syndrome” OR AB “severe acute respiratory syndrome” OR TI “severe acute respiratory syndromes” OR AB “severe acute respiratory syndromes” OR TI sars OR AB sars OR (MH “respiratory syncytial viruses+”) OR TI “respiratory syncytial virus” OR AB “respiratory syncytial virus” OR TI “respiratory syncytial viruses” OR AB “respiratory syncytial viruses” OR TI rsv OR AB rsv OR TI parainfluenza OR AB parainfluenza OR ((Transmission) AND (Coughing OR Sneezing)) OR ((TI respiratory OR AB respiratory AND Tract) AND (TI infection OR AB infection OR TI Infections OR AB Infections OR TI illness OR AB illness)))

AND

((MH “Handwashing+”) OR TI handwashing OR AB handwashing OR TI hand-washing OR AB hand-washing OR ((TI Hand OR AB Hand OR TI Alcohol OR AB Alcohol) AND (TI wash OR AB wash OR TI Washing OR AB Washing OR TI Cleansing OR AB Cleansing OR TI Rinses OR AB Rinses OR TI hygiene OR AB hygiene OR TI rub OR AB rub OR TI Rubbing OR AB Rubbing OR TI sanitiser OR AB sanitiser OR

TI sanitizer OR AB sanitizer OR TI cleanser OR AB cleanser OR TI disinfected OR AB disinfected OR TI Disinfectant OR AB Disinfectant OR TI Disinfect OR AB Disinfect OR TI antiseptic OR AB antiseptic OR TI virucid OR AB virucid)) OR (MH “gloves+”) OR TI Glove OR AB Glove OR Gloves OR (MH “Masks+”) OR (MH “respiratory protective devices+”) OR TI facemask OR AB facemask OR TI Facemasks OR AB Facemasks OR TI mask OR AB mask OR TI Masks OR AB Masks OR TI respirator OR AB respirator OR TI respirators OR AB respirators OR (MH “Protective Clothing”) OR (MH “Protective Devices+”) OR TI “patient isolation” OR AB “patient isolation” OR ((TI school OR AB school OR TI Schools OR AB Schools) AND (TI Closure OR AB Closure OR TI Closures OR AB Closures OR TI Closed OR AB Closed)) OR (MH “Quarantine+”) OR TI quarantine OR AB quarantine OR TI “Hygiene intervention” OR AB “Hygiene intervention” OR (MH “Mouthwashes+”) OR TI gargling OR AB gargling OR TI “nasal tissues” OR AB “nasal tissues”)

AND

((MH “Infection Control+”) OR (MH “Disease Outbreaks+”) OR (MH “Infection Control+”) OR TI “Communicable Disease Control” OR AB “Communicable Disease Control” OR TI “Secondary transmission” OR AB “Secondary transmission” OR ((TI Reduced OR AB Reduced OR TI Reduce OR AB Reduce OR TI Reduction OR AB Reduction OR TI Reducing OR AB Reducing OR TI Lower OR AB Lower) AND (TI Incidence OR AB Incidence OR TI Occurrence OR AB Occurrence OR TI Transmission OR AB Transmission OR TI Secondary OR AB Secondary)))

AND

((MH “Clinical Trials+”) OR (MH “Quantitative Studies”) OR TI placebo* OR AB placebo* OR (MH “Placebos”) OR (MH “Random Assignment”) OR TI random* OR AB random* OR TI ((singl* or doubl* or tripl* or trebl*) W1 (blind* or mask*)) OR AB ((singl* or doubl* or tripl* or trebl*) W1 (blind* or mask*)) OR TI clinic* trial* OR AB clinic* trial* OR PT clinical trial)

## References

1. Forum of International Respiratory Societies. The global impact of respiratory disease, 2017. Available from: https://www.who.int/gard/publications/The_Global_Impact_of_Respiratory_Disease.pdf

2. Bonn D. Spared an influenza pandemic for another year? Lancet (London, England) 1997;349(9044):36–36. doi: 10.1016/s0140-6736(05)62175-7

3. Jefferson T, Demicheli V, Rivetti D, et al. Antivirals for influenza in healthy adults: systematic review. Lancet (London, England) 2006;367(9507):303–13. doi: 10.1016/s0140-6736(06)67970-1

4. Thomas RE, Jefferson T, Lasserson TJ. Influenza vaccination for healthcare workers who work with the elderly. Cochrane Database of Systematic Reviews 2010(2) doi: 10.1002/14651858.CD005187.pub3

5. Jefferson T, Jones MA, Doshi P, et al. Neuraminidase inhibitors for preventing and treating influenza in healthy adults and children. Cochrane Database Syst Rev 2014(4):CD008965. doi: 10.1002/14651858.CD008965.pub4 [published Online First: 2014/04/11]

6. World Health Organisation. Global Influenza Strategy 2019 – 2030. 2019

7. Jefferson T, Del Mar C, Dooley L, et al. Physical interventions to interrupt or reduce the spread of respiratory viruses: systematic review. Bmj 2009;339:b3675. doi: 10.1136/bmj.b3675 [published Online First: 2009/09/24]

8. Jefferson T, Del Mar CB, Dooley L, et al. Physical interventions to interrupt or reduce the spread of respiratory viruses. Cochrane Database Syst Rev 2011(7):Cd006207. doi: 10.1002/14651858.CD006207.pub4 [published Online First: 2011/07/08]

9. Jefferson T, Foxlee R, Del Mar C, et al. Interventions for the interruption or reduction of the spread of respiratory viruses. Cochrane Database of Systematic Reviews 2007(4) doi: 10.1002/14651858.CD006207.pub2

10. Clark J, Glasziou P, Del Mar C, et al. A full systematic review was completed in 2 weeks using automation tools: a case study. J Clin Epidemiol 2020;121:81–90. doi: 10.1016/j.jclinepi.2020.01.008 [published Online First: 2020/02/01]

11. Clark J, Sanders S, Carter M, et al. Improving the translation of search strategies using the Polyglot Search Translator: a randomised controlled trial. J Am Libr Assoc 2020;In press

12. Marshall IJ, Noel-Storr A, Kuiper J, et al. Machine learning for identifying Randomized Controlled Trials: An evaluation and practitioner’s guide. Research synthesis methods 2018;9(4):602–14. doi: 10.1002/jrsm.1287 [published Online First: 2018/01/10]

13. Higgins JP, Altman DG, Gotzsche PC, et al. The Cochrane Collaboration’s tool for assessing risk of bias in randomised trials. BMJ 2011;343:d5928. doi: 10.1136/bmj.d5928 [published Online First: 2011/10/20]

14. Hoffmann TC, Glasziou PP, Boutron I, et al. Better reporting of interventions: template for intervention description and replication (TIDieR) checklist and guide. Bmj 2014;348:g1687. doi: 10.1136/bmj.g1687 [published Online First: 2014/03/13]

15. Higgins JPT, Thompson SG. Quantifying heterogeneity in a meta-analysis. Statistics in Medicine 2002;21(11):1539–58. doi: 10.1002/sim.1186

16. Aiello AE, Coulborn RM, Perez V, et al. A randomized intervention trial of mask use and hand hygiene to reduce seasonal influenza-like illness and influenza infections among young adults in a university setting. International Journal of Infectious Diseases 2010;14:E320–E20. doi: 10.1016/j.ijid.2010.02.2201

17. Cowling BJ, Fung ROP, Cheng CKY, et al. Preliminary Findings of a Randomized Trial of Non-Pharmaceutical Interventions to Prevent Influenza Transmission in Households. Plos One 2008;3(5) doi: 10.1371/journal.pone.0002101

18. Jacobs JL, Ohde S, Takahashi O, et al. Use of surgical face masks to reduce the incidence of the common cold among health care workers in Japan: A randomized controlled trial. American journal of infection control 2009;37(5):417–19. doi: 10.1016/j.ajic.2008.11.002

19. Loeb M, Dafoe N, Mahony J, et al. Surgical mask vs N95 respirator for preventing influenza among health care workers: a randomized trial. Jama 2009;302(17):1865–71. doi: 10.1001/jama.2009.1466 [published Online First: 2009/10/03]

20. MacIntyre CR, Cauchemez S, Dwyer DE, et al. Face mask use and control of respiratory virus transmission in households. Emerging infectious diseases 2009;15(2):233–41. doi: 10.3201/eid1502.081167 [published Online First: 2009/02/06]

21. Suess T, Remschmidt C, Schink SB, et al. The role of facemasks and hand hygiene in the prevention of influenza transmission in households: results from a cluster randomised trial; Berlin, Germany, 2009-2011. BMC Infect Dis 2012;12:26. doi: 10.1186/1471-2334-12-26. [published Online First: 2012/01/28]

22. Aiello AE, Perez V, Coulborn RM, et al. Facemasks, hand hygiene, and influenza among young adults: a randomized intervention trial. PLoS One 2012;7(1):e29744. doi: 10.1371/journal.pone.0029744. Epub 2012 Jan 25. [published Online First: 2012/02/02]

23. Barasheed O, Almasri N, Badahdah AM, et al. Pilot Randomised Controlled Trial to Test Effectiveness of Facemasks in Preventing Influenza-like Illness Transmission among Australian Hajj Pilgrims in 2011. Infect Disord Drug Targets 2014;14(2):110–6. doi: 10.2174/1871526514666141021112855 [published Online First: 2014/10/23]

24. Canini L, Andreoletti L, Ferrari P, et al. Surgical mask to prevent influenza transmission in households: a cluster randomized trial. PLoS One 2010;5(11):e13998. doi: 10.1371/journal.pone.0013998. [published Online First: 2010/11/26]

25. MacIntyre CR, Seale H, Dung TC, et al. A cluster randomised trial of cloth masks compared with medical masks in healthcare workers. BMJ Open 2015;5(4):e006577. doi: 10.1136/bmjopen-2014-006577. [published Online First: 2015/04/24]

26. MacIntyre CR, Zhang Y, Chughtai AA, et al. Cluster randomised controlled trial to examine medical mask use as source control for people with respiratory illness. BMJ Open 2016;6(12):e012330. doi: 10.1136/bmjopen-2016-012330. [published Online First: 2017/01/01]

27. Loeb M, Dafoe N, Mahony J, et al. Surgical Mask vs N95 Respirator for Preventing Influenza Among Health Care Workers A Randomized Trial. Jama-Journal of the American Medical Association 2009;302(17):1865–71. doi: 10.1001/jama.2009.1466

28. MacIntyre CR, Wang Q, Cauchemez S, et al. A cluster randomized clinical trial comparing fit-tested and non-fit-tested N95 respirators to medical masks to prevent respiratory virus infection in health care workers. Influenza Other Respir Viruses 2011;5(3):170–9. doi: 10.1111/j.1750-2659.2011.00198.x. Epub 2011 Jan 27. [published Online First: 2011/04/12]

29. MacIntyre CR, Wang Q, Seale H, et al. A randomized clinical trial of three options for N95 respirators and medical masks in health workers. Am J Respir Crit Care Med 2013;187(9):960–6. doi: 10.1164/rccm.201207-1164OC. [published Online First: 2013/02/16]

30. Radonovich LJ, Jr., Simberkoff MS, Bessesen MT, et al. N95 Respirators vs Medical Masks for Preventing Influenza Among Health Care Personnel: A Randomized Clinical Trial. Jama 2019;322(9):824–33. doi: 10.1001/jama.2019.11645. [published Online First: 2019/09/04]

31. Miyaki K, Sakurazawa H, Mikurube H, et al. An effective quarantine measure reduced the total incidence of influenza A H1N1 in the workplace: another way to control the H1N1 flu pandemic. J Occup Health 2011;53(4):287–92. doi: 10.1539/joh.10-0024-fs. Epub 2011 May 18. [published Online First: 2011/05/21]

32. Elflein J. Estimated number of influenza cases in the United States from 2010 to 2017, 2019.

33. Smith JD, MacDougall CC, Johnstone J, et al. Effectiveness of N95 respirators versus surgical masks in protecting health care workers from acute respiratory infection: a systematic review and meta-analysis. CMAJ 2016;188(8):567–74. doi: 10.1503/cmaj.150835 [published Online First: 2016/03/10]

34. Offeddu V, Yung CF, Low MSF, et al. Effectiveness of Masks and Respirators Against Respiratory Infections in Healthcare Workers: A Systematic Review and Meta-Analysis. Clin Infect Dis 2017;65(11):1934–42. doi: 10.1093/cid/cix681 [published Online First: 2017/11/16]

35. Long Y, Hu T, Liu L, et al. Effectiveness of N95 respirators versus surgical masks against influenza: A systematic review and meta-analysis. J Evid Based Med 2020 doi: 10.1111/jebm.12381 [published Online First: 2020/03/14]

36. World Health Organization. Report of the WHO-China Joint Mission on Coronavirus Disease 2019 (COVID-19) 16-24 February 2020 [Internet]. Geneva World Health Organization, 2020.

37. Chan JFW YS, Kok KH, To KK, Chu H, Yang J, et al. A familial cluster of pneumonia associated with the 2019 novel coronavirus indicating person-to-person transmission: a study of a family cluster. Lancet Infectious Diseases 2020;395(10223):514–23.

38. World Health Organozation. Infection prevention and control of epidemic-and pandemic-prone acute respiratory infections in health care., 2014.

39. Zhu JH LS, Wang DY, et al. J Lung Pulm Respir Res. Effects of long-duration wearing of N95 respirator and surgical facemask: a pilot study. 2014;1(14):97–100.

40. Rebmann T CR, Wang J. Physiologic and other effects and compliance with long-term respirator use among medical intensive care unit nurses. Am J Infect Control 2013 41(12):1218–23.

41. Foo CCI, Goon ATJ, Leow Y-H, et al. Adverse skin reactions to personal protective equipment against severe acute respiratory syndrome - a descriptive study in Singapore. Contact Dermatitis 2006;55(5):291–94. doi: 10.1111/j.1600-0536.2006.00953.x

42. Tan KT GM. N95 acne. Int J Dermatol 2004;43(7):522–23.

43. Donovan J KI, Holness LD, Skotnicki-Grant S, et al. Skin reactions following use of N95 facial masks. Dermatitis 2007;18(2):104.;18(2):104.

44. Donovan J S-GS. Allergic contact dermatitis from formaldehyde textile resins in surgical uniforms and nonwoven textile masks. Dermatitis 2007;18(1):40–44.

